# Genetically-proxied therapeutic inhibition of antihypertensive drug targets and risk of common cancers

**DOI:** 10.1101/2021.03.11.21252971

**Authors:** James Yarmolinsky, Virginia Díez-Obrero, Tom G Richardson, Marie Pigeyre, Jennifer Sjaarda, Guillaume Paré, Venexia M Walker, Emma E Vincent, Vanessa Y Tan, Mireia Obón-Santacana, Demetrius Albanes, Jochen Hampe, Andrea Gsur, Heather Hampel, Ellen Kampman, Rish K Pai, Mark Jenkins, Steven Gallinger, Graham Casey, Wei Zheng, Christopher I Amos, the International Lung Cancer Consortium, the PRACTICAL consortium, George Davey Smith, Richard M Martin, Victor Moreno

## Abstract

**Background:** Epidemiological studies have reported conflicting findings on the potential adverse effects of long-term antihypertensive medication use on cancer risk. Naturally occurring variation in genes encoding antihypertensive drug targets can be used as proxies for these targets to examine the effect of their long-term therapeutic inhibition on disease outcomes.

**Methods:** Single-nucleotide polymorphisms (SNPs) in *ACE*, *ADRB1*, and *SLC12A3* associated (*P* < 5.0 x 10^-8^) with systolic blood pressure in genome-wide association studies (GWAS) were used to proxy inhibition of angiotensin-converting enzyme (ACE), β-1 adrenergic receptor (ADRB1), and sodium-chloride symporter (NCC), respectively. Summary genetic association estimates for these SNPs were obtained from GWAS consortia for the following cancers: breast (122,977 cases, 105,974 controls), colorectal (58,221 cases, 67,694 controls), lung (29,266 cases, 56,450 controls), and prostate (79,148 cases, 61,106 controls). Replication analyses were performed in the FinnGen consortium (1,573 colorectal cancer cases, 120,006 controls). Inverse-variance weighted random-effects models were used to examine associations between genetically-proxied inhibition of these drug targets and risk of cancer. Multivariable Mendelian randomization and colocalisation analyses were employed to examine robustness of findings to violations of Mendelian randomization assumptions.

**Results:** Genetically-proxied ACE inhibition equivalent to a 1 mmHg reduction in systolic blood pressure was associated with increased odds of colorectal cancer (OR 1.13, 95% CI 1.06-1.22; *P* = 3.6 x 10^-4^). This finding was replicated in the FinnGen consortium (OR 1.40, 95% CI 1.02-1.92; *P* = 0.035). There was little evidence of association of genetically-proxied ACE inhibition with risk of breast cancer (OR 0.98, 95% CI 0.94-1.02, *P* = 0.35), lung cancer (OR 1.01, 95% CI 0.92-1.10; *P* = 0.93), or prostate cancer (OR 1.06, 95% CI 0.99-1.13; *P* = 0.08). Genetically-proxied inhibition of ADRB1 and NCC were not associated with risk of these cancers.

**Conclusion:** Genetically-proxied long-term ACE inhibition was associated with an increased risk of colorectal cancer, warranting comprehensive evaluation of the safety profiles of ACE inhibitors in clinical trials with adequate follow-up. There was little evidence to support associations across other drug target-cancer risk analyses, consistent with findings from short-term randomised controlled trials for these medications.

## Background

Angiotensin-converting enzyme (ACE) inhibitors are commonly prescribed antihypertensive medications[1]. These medications lower blood pressure by inhibiting the conversion of angiotensin I to angiotensin II, a vasoconstrictor and the primary effector molecule of the renin-angiotensin system (RAS). Though clinical trials have supported the relative safety of these medications in the short-term (median follow-up of 3.5 years), concerns have been raised that long-term use of these medications could increase risk of cancer [2, 3]. These safety concerns relate to the multifaceted role of ACE which cleaves various other substrates beyond angiotensin I, including several peptides that have proliferative effects. For example, ACE inhibition leads to the accumulation of bradykinin, an inflammatory mediator involved in tumour growth and metastasis[4]. In addition, substance P is elevated in ACE inhibitor users which can promote tumour proliferation, migration, and angiogenesis [5, 6].

Some observational epidemiological studies have suggested potential adverse effects of long-term use of these drugs on risk of common cancers (i.e. breast, colorectal, lung, and prostate) [7–10], though findings have been largely inconsistent [11–13]. Interpretation of the epidemiological literature is challenging for several reasons. First, pharmaco-epidemiological studies are susceptible to residual confounding due to unmeasured or imprecisely measured confounders, including those related to indication[14]. Second, several studies examining ACE inhibitor use and cancer risk have included prevalent drug users which can introduce bias because prevalent users are “survivors” of the early period of pharmacotherapy and because covariates at study entry can be influenced by prior medication use[12, 15–18]. Third, some prior studies may have suffered from time-related biases, including immortal time bias which can arise because of misalignment of the start of follow-up, eligibility, and treatment assignment of participants[15, 16, 18, 19]. These biases can produce illusory results in favour of the treatment group while other biases often pervasive in the pharmaco- epidemiological literature (e.g. detection bias due to more intensive clinical monitoring and testing of individuals receiving treatment) can alternatively generate upward-biased effect estimates among those receiving treatment.

Along with ACE inhibitors, β blockers and thiazide diuretics are commonly prescribed antihypertensive medications that lower blood pressure through pathways independent to that of ACE (i.e. β blockers bind to β-adrenergic receptors, inhibiting the binding of norepinephrine and epinephrine to these receptors; thiazide diuretics promote sodium and water excretion by inhibiting sodium reabsorption in renal tubules)[4]. In contrast to ACE inhibitors, some *in vitro* and epidemiological studies have suggested potential chemo-preventive effects of these medications on cancer risk though findings have been inconclusive [20–28].

Naturally occurring variation in genes encoding antihypertensive drug targets can be used as proxies for these targets to examine the effect of their therapeutic inhibition on disease outcomes (“Mendelian randomization”) [29, 30]. Such an approach should be less prone to conventional issues of confounding as germline genetic variants are randomly assorted at meiosis. In addition, Mendelian randomization analysis permits the effect of long-term modulation of drug targets on cancer risk to be examined. Drug-target Mendelian randomization can therefore be used to mimic the effect of pharmacologically modulating a drug target in clinical trials and has been used previously to anticipate clinical benefits and adverse effects of therapeutic interventions[31–34].

We used a Mendelian randomization approach to examine the effect of long-term inhibition of the drug targets for ACE inhibitors (ACE; angiotensin-converting enzyme), β blockers (ADRB1; beta-1 adrenergic receptor), and thiazide diuretic agents (NCC; sodium-chloride symporter) on risk of overall and subtype-specific breast, colorectal, lung, and prostate cancer.

## Methods

### Study populations

For primary analyses, summary genetic association data were obtained from four cancer genome-wide association study (GWAS) consortia. Summary genetic association estimates for overall and oestrogen receptor-stratified breast cancer risk in up to 122,977 cases and 105,974 controls were obtained from the Breast Cancer Association Consortium (BCAC)[35]. Summary genetic association estimates for overall and site-specific colorectal cancer risk in up to 58,221 cases and 67,694 controls were obtained from an analysis of the Genetics and Epidemiology of Colorectal Cancer Consortium (GECCO), ColoRectal Transdisciplinary Study (CORECT), and Colon Cancer Family Registry (CCFR)[36]. Summary genetic association estimates for overall and histological subtype-stratified lung cancer risk in up to 29,266 cases and 56,450 controls were obtained from an analysis of the Integrative Analysis of Lung Cancer Risk and Etiology (INTEGRAL) team of the International Lung Cancer Consortium (ILCCO) [37]. Summary genetic association estimates for overall and advanced prostate cancer risk in up to 79,148 cases and 61,106 controls were obtained from the Prostate Cancer Association Group to Investigate Cancer Associated Alterations in the Genome (PRACTICAL) consortium[38]. These analyses were restricted to participants of European ancestry.

For replication analyses, summary genetic association data were obtained on 1,573 colorectal cancer cases and 120,006 controls of European ancestry from the Finngen consortium. We also examined whether findings could be extended to individuals of East Asian ancestry by obtaining summary genetic association data on 23,572 colorectal cancer cases and 48,700 controls of East Asian ancestry from a GWAS meta-analysis of the Asia Colorectal Cancer Consortium and the Korean National Cancer Center CRC Study 2[39].

Further information on statistical analysis, imputation, and quality control measures for these studies is available in the original publications. All studies contributing data to these analyses had the relevant institutional review board approval from each country, in accordance with the Declaration of Helsinki, and all participants provided informed consent.

### Instrument construction

To generate instruments to proxy ACE, ADRB1, and NCC inhibition, we pooled summary genetic association data from two previously published GWAS of systolic blood pressure (SBP) using inverse-variance weighted fixed-effects models in METAL[40]. The first GWAS was a meta-analysis of ≤ 757,601 individuals of European descent in the UK Biobank and International Consortium of Blood Pressure-Genome Wide Association Studies (ICBP)[41]. The second GWAS was performed in 99,785 individuals in the Genetic Epidemiology Research on Adult Health and Aging (GERA) cohort, of whom the majority (81.0%) were of European ancestry[42]. Both GWAS were adjusted for age, sex, body mass index (BMI), and antihypertensive medication use. Estimates that were genome-wide significant (*P* < 5.0 x 10^-8^) in pooled analyses (N ≤ 857,386) and that showed concordant direction of ≤ effect across both GWAS were then used to generate instruments.

To proxy ADRB1 inhibition, 8 SNPs associated with systolic blood pressure at genome-wide significance and within ± 100 kb windows from *ADRB1* were obtained. To proxy NCC inhibition, 1 SNP associated with systolic blood pressure at genome-wide significance and within a ± 100 kb window from *SLC12A3* (alias for *NCC*) was obtained. For both of these drug targets, SNPs used as proxies were permitted to be in weak linkage disequilibrium (r^2^ < 0.10) with each other to increase the proportion of variance in each respective drug target explained by the instrument, maximising instrument strength.

Since pooled GWAS estimates were obtained from analyses adjusted for BMI, which could induce collider bias, we also examined constructing instruments using summary genetic association data from a previous GWAS of systolic blood pressure in 340,159 individuals in UK Biobank without adjustment for BMI or antihypertensive medication use (**Supplementary Table 1**) [43].

We explored construction of genetic instruments to proxy ACE inhibition using two approaches: i) by obtaining genome-wide significant variants in weak linkage disequilibrium (r^2^ < 0.10) in or within ± 100 kb from *ACE* that were associated with systolic blood pressure in previously described pooled GWAS analyses (resulting in two SNPs) and ii) by obtaining genome-wide significant variants in weak linkage disequilibrium (r^2^ < 0.10) in or within ± 100 kb from *ACE* that were associated with serum ACE concentrations in a GWAS of 4,174 participants in the Outcome Reduction with Initial Glargine INtervention (ORIGIN) study (resulting in 14 SNPs) [44]. 46.6% of participants in the ORIGIN study were of European ancestry and 53.4% were of Latin American ancestry. Effect allele frequencies for these 14 SNPs were broadly similar across both ancestries (**Supplementary Table 2**). We then compared the proportion of variance in either systolic blood pressure or serum ACE concentrations explained (r^2^) across each respective instrument to prioritise the primary instrument to proxy ACE inhibition. The serum ACE concentrations instrument (r^2^ = 0.34-0.39, F= 2,156.5-2,594.9) was prioritised because of stronger instrument strength as compared to the systolic blood pressure instrument (r^2^ =0.02, F=128.5).

To validate the serum ACE concentrations instrument, we examined the association between genetically-proxied ACE inhibition and i) systolic blood pressure, ii) risk of stroke in the MEGASTROKE consortium (40,585 cases; 406,111 controls of European ancestry), iii) risk of coronary artery disease in the CARDIoGRAMplusC4D consortium (60,801 cases; 123,504 controls, 77% of whom were of European ancestry), and iv) risk of type 2 diabetes in the DIAGRAM consortium (N=74,124 cases; 824,006 controls of European ancestry) and compared the direction of effect estimates obtained with those reported for ACE inhibitor use in meta-analyses of randomised controlled trials[45–47]. Likewise, we validated ADRB1 and NCC instruments by examining the association between inhibition of these targets and risk of stroke and coronary artery disease, as reported in meta-analyses of clinical trials[47].

For analyses in individuals of East Asian ancestry, 1 *cis*-acting variant (rs4343) associated with ACE activity (*P* = 3.0 x 10^-25^) in a GWAS of 623 individuals with young onset hypertension of Han Chinese descent was obtained[48]. In the Japanese Biobank (N=136,597), the A allele of rs4343 has previously been shown to associate with lower SBP (-0.26 mmHg SBP, 95% CI -0.11 to -0.42; *P* = 6.7 x 10^-4^)[49]. This variant explained 0.008% of the variance of SBP (F=11.6).

### Mendelian randomization primary and sensitivity analyses

Inverse-variance weighted random-effects models were employed to estimate causal effects of genetically-proxied drug target inhibition on cancer risk. These models were adjusted for weak linkage disequilibrium between SNPs (r^2^ < 0.10) with reference to the 1000 Genomes Phase 3 reference panel[50, 51]. If under-dispersion in causal estimates generated from individual genetic variants was present, the residual standard error was set to 1.

Mendelian randomization analysis assumes that the genetic instrument used to proxy a drug target (i) is associated with the drug target (“relevance”), (ii) does not share a common cause with the outcome (“exchangeability”), and (iii) affects the outcome only through the drug target (“exclusion restriction”).

We tested the “relevance” assumption by generating estimates of the proportion of variance of each drug target explained by the instrument (r^2^) and F-statistics. F-statistics can be used to examine whether results are likely to be influenced by weak instrument bias, i.e., reduced statistical power when an instrument explains a limited proportion of the variance in a drug target. As a convention, an F-statistic of at least 10 is indicative of minimal weak instrument bias[52].

We evaluated the “exclusion restriction” assumption by performing various sensitivity analyses. First, we performed colocalisation to examine whether drug targets and cancer endpoints showing nominal evidence of an association in MR analyses (*P* < 0.05) share the same causal variant at a given locus. Such an analysis can permit exploration of whether drug targets and cancer outcomes are influenced by distinct causal variants that are in linkage disequilibrium with each other, indicative of horizontal pleiotropy (an instrument influencing an outcome through pathways independent to that of the exposure), a violation of the exclusion restriction criterion [53]. Colocalisation analysis was performed by generating ± 300 kb windows from the top SNP used to proxy each respective drug target. As a convention, a posterior probability of ≥ 0.80 was used to indicate support for a configuration tested. An extended description of colocalisation analysis including assumptions of this method is presented in **Supplementary Material**.

For analyses showing evidence of colocalisation across drug target and cancer endpoint signals, we then examined whether there was evidence of an association of genetically-proxied inhibition of that target with previously reported risk factors for the relevant cancer endpoint (e.g. body mass index, low-density lipoprotein cholesterol, total cholesterol, iron, and insulin-like growth factor 1 for colorectal cancer risk) [54–60]. If there was evidence for an association between a genetically-proxied drug target and previously reported risk factor (*P* < 0.05), which could suggest the presence of horizontal pleiotropy, multivariable Mendelian randomization can then be used to examine the association of drug target inhibition in relation to cancer risk, accounting for this risk factor[61].

Finally, iterative leave-one-out analysis was performed iteratively removing one SNP at a time from instruments to examine whether findings were driven by a single influential SNP.

To account for multiple testing across primary drug target analyses, a Bonferroni correction was used to establish a *P*-value threshold of < 0.0014 (false positive rate = 0.05/36 statistical tests [3 drug targets tested against 12 cancer endpoints]).

### Colon transcriptome-wide Mendelian randomization analysis

To explore potential mechanisms governing associations and to further evaluate potential violations of Mendelian randomization assumptions, we examined associations of genetically-proxied ACE inhibition with gene expression profiles in normal (i.e. non-neoplastic) colon tissue samples. Gene expression analysis was performed using data from the University of Barcelona and the University of Virginia Genotyping and RNA Sequencing Project (BarcUVa-Seq)[62]. This analysis was restricted to 445 individuals (mean age 60 years, 64% female, 95% of European ancestry) who participated in a Spanish colorectal cancer risk screening programme who obtained a normal colonoscopy result (i.e. macroscopically normal colon tissue, with no malignant lesions). Further information on RNA-Seq data processing and quality control is presented in **Supplementary Material**.

To perform transcriptome-wide analyses, weighted genetic risk scores (wGRS) to proxy serum ACE concentrations were constructed using 14 ACE SNPs in Plink v1.9[63]. Expression levels for 21,482 genes (expressed as inverse normal transformed trimmed mean of M-values) were regressed on the standardised wGRS and adjusted for sex, the top two principal components of genetic ancestry, sequencing batch, probabilistic estimation of expression residuals (PEER) factors, and colon anatomical location. To account for multiple testing, a Bonferroni correction was used to establish a *P*-value threshold of < 2.33 x 10^-6^ (false positive rate = 0.05/21,482 statistical tests).

Bioinformatic follow-up of findings from transcriptome-wide analysis was performed to further interrogate downstream perturbations of the ACE wGRS on gene expression profiles using gene-set enrichment analysis and co-expression network analysis. In brief, these methods can either evaluate whether expression levels of genes associated with the ACE wGRS are enriched in relation to an *a priori* defined set of genes based on curated functional annotation (gene-set enrichment analysis) or permit the identification of clusters of genes (termed “modules”) which show a coordinated expression pattern associated with the wGRS (co-expression network analysis). Further information on gene-set enrichment and co-expression network analysis is presented in **Supplementary Material**.

## Results

Across the 3 drug targets that we examined, conservative estimates of F-statistics for their respective genetic instruments ranged from 269.1 to 2,156.5, suggesting that our analyses were unlikely to suffer from weak instrument bias. Characteristics of genetic variants in *ACE*, *ADRB1*, and *SLC12A3* used to proxy each pharmacological target are presented in **Table 1**. Estimates of r^2^ and F-statistics for each target are presented in **Supplementary Table 3**.

**Table 1.** Characteristics of systolic blood pressure lowering genetic variants in *ACE*, *ADRB1*, and *SLC12A3*

| Target | Effect allele/Non-Effect Allele | Effect Allele Frequency | Effect (SE) | P-value |
| --- | --- | --- | --- | --- |
| *ACE* |  |  |  |  |
| rs4343 | A/G | 0.45 | -0.63 (0.02) | $1.53 \times 10^{-213}$ |
| rs12452187 | A/G | 0.60 | -0.23 (0.02) | $2.53 \times 10^{-27}$ |
| rs79480822 | C/T | 0.93 | -0.55 (0.05) | $6.37 \times 10^{-24}$ |
| rs3730025 | G/A | 0.01 | -0.80 (0.09) | $4.32 \times 10^{-19}$ |
| rs11655956 | C/G | 0.08 | -0.35 (0.04) | $1.06 \times 10^{-15}$ |
| rs118121655 | G/A | 0.96 | -0.54 (0.07) | $3.10 \times 10^{-15}$ |
| rs4365 | G/A | 0.97 | -0.58 (0.08) | $7.06 \times 10^{-12}$ |
| rs4968771 | G/A | 0.08 | -0.22 (0.03) | $1.78 \times 10^{-11}$ |
| rs12150648 | G/A | 0.96 | -0.39 (0.06) | $1.88 \times 10^{-10}$ |
| rs80311894 | T/G | 0.97 | -0.46 (0.07) | $2.60 \times 10^{-10}$ |
| rs118138685 | C/G | 0.04 | -0.40 (0.07) | $2.44 \times 10^{-9}$ |
| rs13342595 | C/T | 0.23 | -0.14 (0.02) | $2.48 \times 10^{-9}$ |
| rs28656895 | T/C | 0.23 | -0.14 (0.02) | $3.77 \times 10^{-9}$ |
| rs4968780 | C/A | 0.05 | -0.28 (0.05) | $1.86 \times 10^{-8}$ |
| *ADRB1* |  |  |  |  |
| rs1801253 | G/C | 0.23 | -0.41 (0.03) | $8.07 \times 10^{-43}$ |
| rs11196549 | G/A | 0.96 | -0.62 (0.07) | $2.53 \times 10^{-19}$ |
| rs4918889 | G/C | 0.17 | -0.30 (0.04) | $7.53 \times 10^{-18}$ |
| rs460718 | A/G | 0.33 | -0.24 (0.03) | $2.21 \times 10^{-17}$ |
| rs11196597 | G/A | 0.86 | -0.27 (0.04) | $3.07 \times 10^{-12}$ |
| rs143854972 | G/A | 0.94 | -0.39 (0.06) | $4.35 \times 10^{-11}$ |
| rs17875473 | C/T | 0.91 | -0.28 (0.05) | $9.04 \times 10^{-9}$ |
| rs10787510 | A/G | 0.48 | -0.15 (0.03) | $2.01 \times 10^{-8}$ |
| *NCC* |  |  |  |  |
| rs35797045 | A/C | 0.05 | -0.35 (0.06) | $4.85 \times 10^{-8}$ |

Effect (SE) represents change in serum ACE concentrations per additional copy of the effect allele for ACE analysis and change in systolic blood pressure per additional copy of the effect allele for ADRB1 and NCC analyses. In analyses of genetically-proxied ACE inhibition and colorectal cancer risk, one SNP (rs8064760) was not available in the colorectal cancer dataset. Two SNPs associated with systolic blood pressure used to proxy ACE inhibition in sensitivity analyses were: rs8077276 (effect allele/non-effect allele: A/G, effect (se): -0.27 (0.03), effect allele frequency: 0.62; *P*-value: $4.47 \times 10^{-22}$ ) and rs28656895 (effect allele/non-effect allele: T/C, effect (se): -0.19 (0.03), effect allele frequency: 0.23; *P*-value: $3.37 \times 10^{-9}$ ).

### Instrument validation

Findings from genetic instrument validation analyses for drug targets were broadly concordant (i.e. in direction of effect) with findings from meta-analyses of randomised trials for these medications.

Genetically-proxied ACE inhibition was associated with lower SBP (mmHg per SD lower serum ACE concentration: -0.40, 95% CI -0.21 to -0.59, *P* = 4.2 x 10^-5^) and a lower risk of type 2 diabetes (OR equivalent to 1 mmHg lower SBP: 0.90, 95% CI 0.85-0.95, *P* = 1.3 x 10^-4^). There was weak evidence for an association of genetically-proxied ACE inhibition with lower risk of stroke (OR 0.94, 95% CI 0.88-1.01; *P* = 0.06) and coronary artery disease (OR 0.95, 95% CI 0.89-1.02; *P* = 0.16).

Genetically-proxied ADRB1 inhibition was associated with lower risk of coronary artery disease (per 1mmHg lower SBP: OR 0.95, 95% CI 0.92-0.98; *P* = 1.5 x 10^-3^) but there was little evidence of association with risk of stroke (OR 1.03, 95% CI 0.99-1.07; *P* = 0.18).

Genetically-proxied NCC inhibition was associated with lower risk of coronary artery disease (per 1mmHg lower SBP: OR 0.81, 95% CI 0.81, 95% CI 0.71-0.93, *P* = 3.2 x 10^-3^) and was weakly associated with lower risk of stroke (OR 0.89, 95% CI 0.78-1.02; *P* = 0.10).

### Genetically-proxied ACE inhibition and cancer risk

Genetically-proxied ACE inhibition was associated with an increased odds of colorectal cancer (OR equivalent to 1 mmHg lower SBP: 1.13, 95% CI 1.06-1.22; *P* = 3.6 x 10^-4^). Likewise, in analyses using SBP SNPs in *ACE*, genetically-proxied SBP lowering via ACE inhibition was associated with an increased odds of colorectal cancer (OR equivalent to 1 mmHg lower SBP: 1.11, 95% CI 1.04-1.18; *P* = 1.3 x 10^-3^). When scaled to represent SBP lowering achieved in clinical trials of ACE inhibitors for primary hypertension (equivalent to 8 mmHg lower SBP), this represents an OR of 2.74 (95% CI 1.58-4.76)[64]. In site-specific analyses, this association was stronger for colon cancer risk (OR 1.18, 95% CI 1.07-1.31; *P* = 9.7 x 10^-4^) than rectal cancer risk (OR 1.07, 95% CI 0.97-1.18; *P* = 0.16).

Similar associations were found across risk of proximal colon cancer (OR 1.23, 95% CI 1.10-1.37; *P* = 1.9 x 10^-4^) and distal colon cancer (OR 1.15, 95% CI 1.03-1.27; *P* = 0.01).

Colocalisation analysis suggested that serum ACE and colorectal cancer associations had a 91.4% posterior probability of sharing a causal variant within the *ACE* locus (**Supplementary Table 4**).

Regional Manhattan plots examining the association of all SNPs ± 300 kb from the top SNP for serum ACE concentrations (rs4343) for their association with serum ACE concentrations (**Figure 1**) and with colorectal cancer risk (**Figure 2**) did not appear to support the presence of two or more independent causal variants driving associations across either trait.

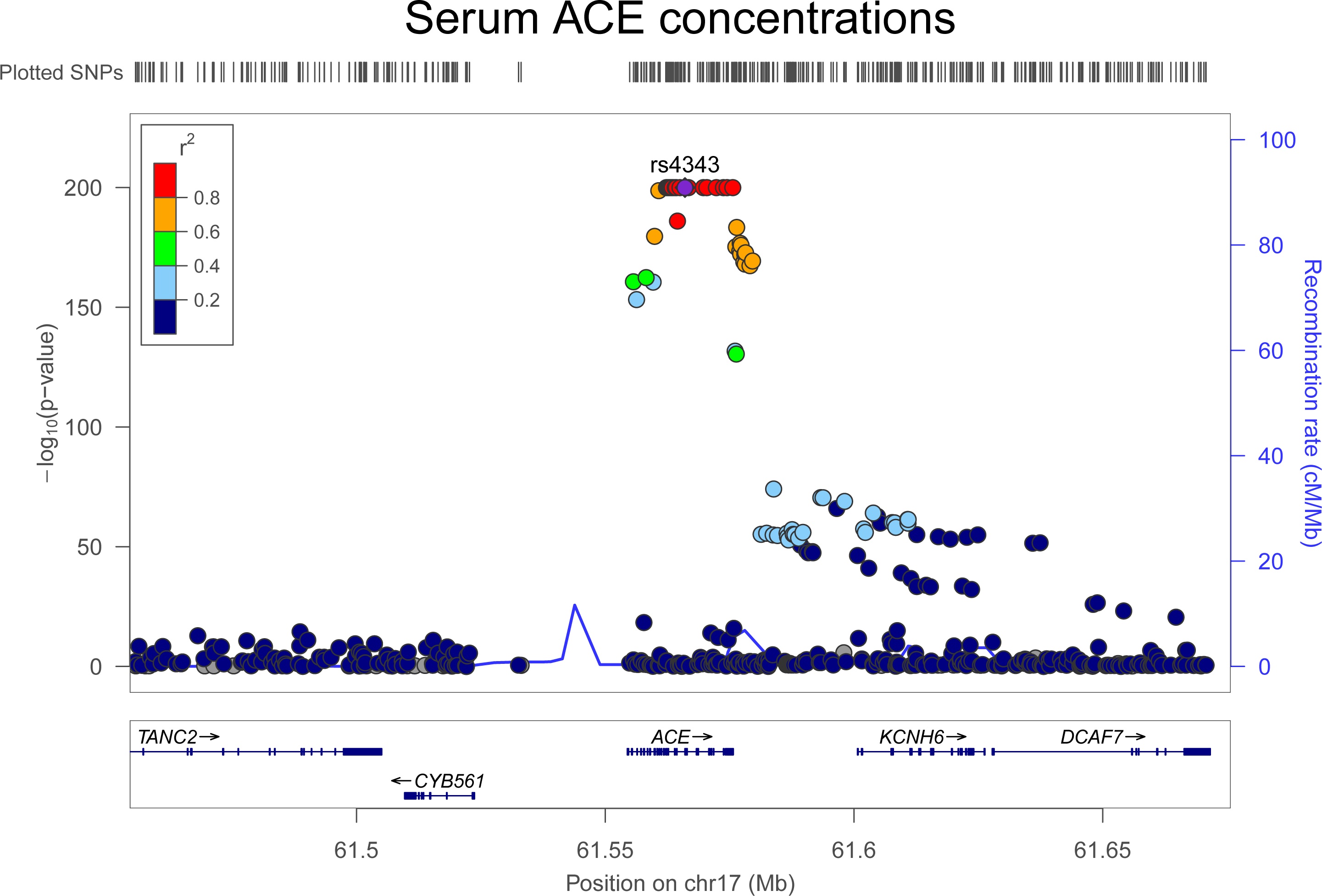

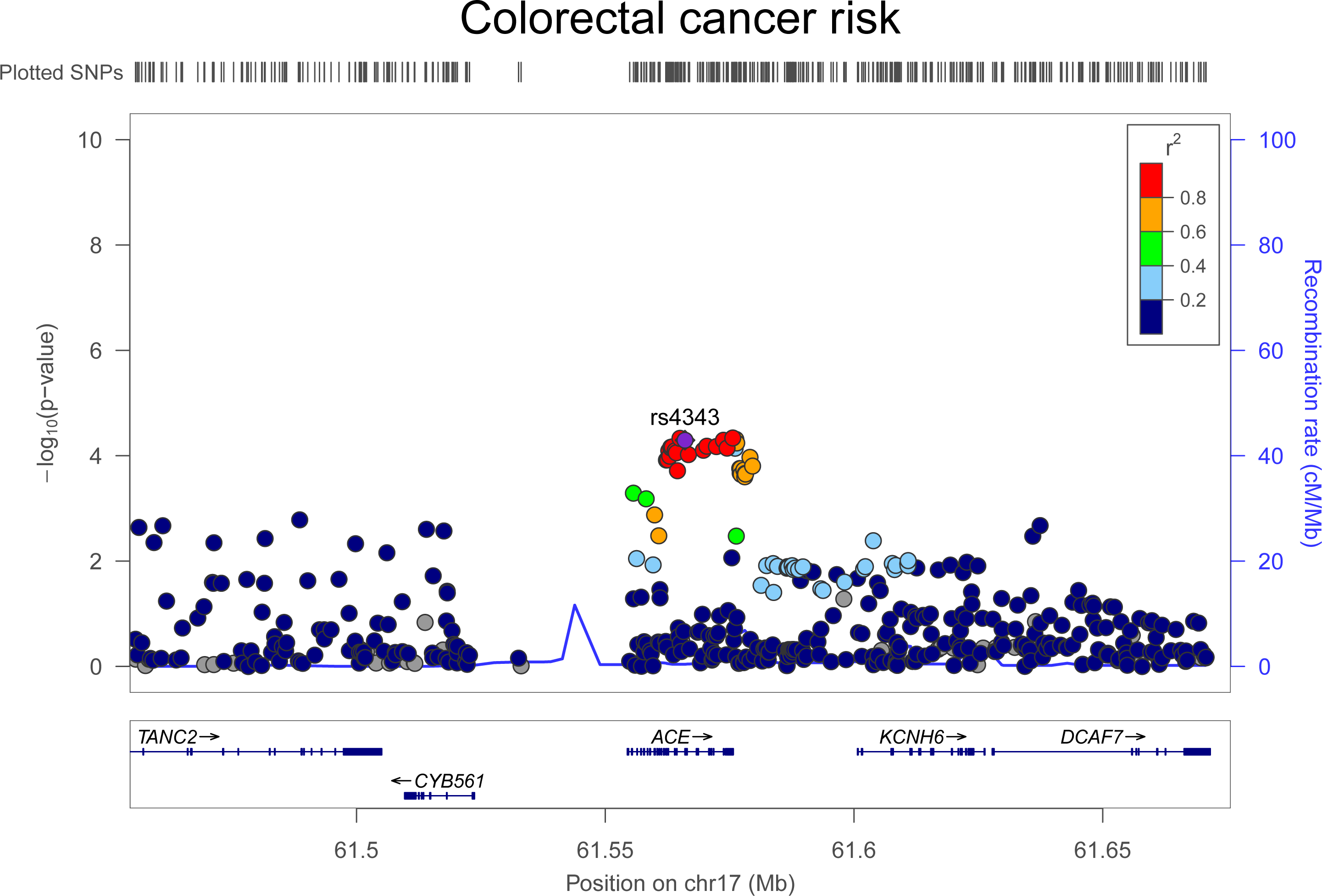

In Mendelian randomization analyses examining the association of genetically-proxied ACE inhibition with previously reported colorectal cancer risk factors, there was little evidence to support associations with body mass index, low-density lipoprotein cholesterol, total serum cholesterol, serum iron, or serum insulin-like growth factor-1 (**Supplementary Table 5**). There was also little evidence to support an association of genetically-proxied systolic blood pressure with colorectal cancer risk (OR per 1mmHg lower SBP: 1.00, 95% CI 0.99-1.01; *P* = 0.50), suggesting a potential mechanism-specific effect of this drug target on colorectal cancer risk.

Additionally, results of analyses that iteratively removed one SNP at a time from the instrument and recalculated the overall Mendelian randomization estimate were consistent, suggesting that associations were not being driven through individual influential SNPs (**Supplementary Table 6**).

There was little evidence that genetically-proxied ACE inhibition was associated with risk of the other cancer sites examined (**Table 2**).

**Table 2.**
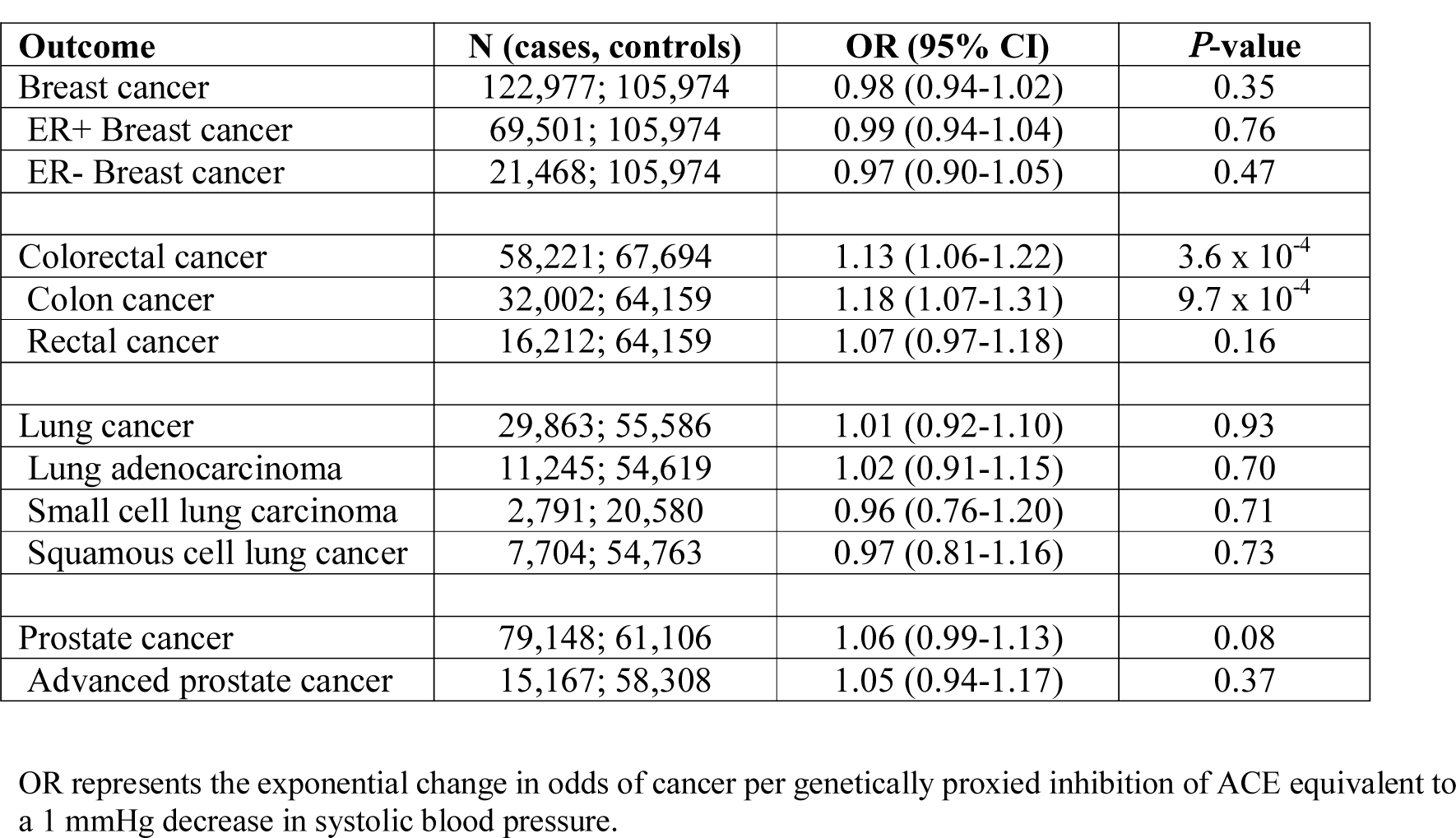
Association between genetically-proxied ACE inhibition and risk of overall and subtypespecific breast, colorectal, prostate, and lung cancer risk

### Genetically-proxied ADRB1 inhibition and cancer risk

There was little evidence that genetically-proxied ADRB1 inhibition was associated with overall risk of breast, colorectal, lung, or prostate cancer (**Table 3**). In lung cancer subtype-stratified analyses, there was evidence to suggest an association of genetically-proxied ADRB1 inhibition with lower risk of small cell lung carcinoma (OR equivalent to 1 mmHg lower SBP: 0.87, 95% CI 0.79-0.96; *P* = 0.008). Colocalisation analysis suggested that ADRB1 and small cell lung carcinoma were unlikely to share a causal variant within the *ADRB1* locus (1.5% posterior probability of a shared causal variant) (**Supplementary Table 7, Figures 3-4**). Findings for overall and subtype-specific cancer risk did not differ markedly when using an instrument for ADRB1 inhibition constructed from a GWAS unadjusted for BMI (**Supplementary Table 8**).

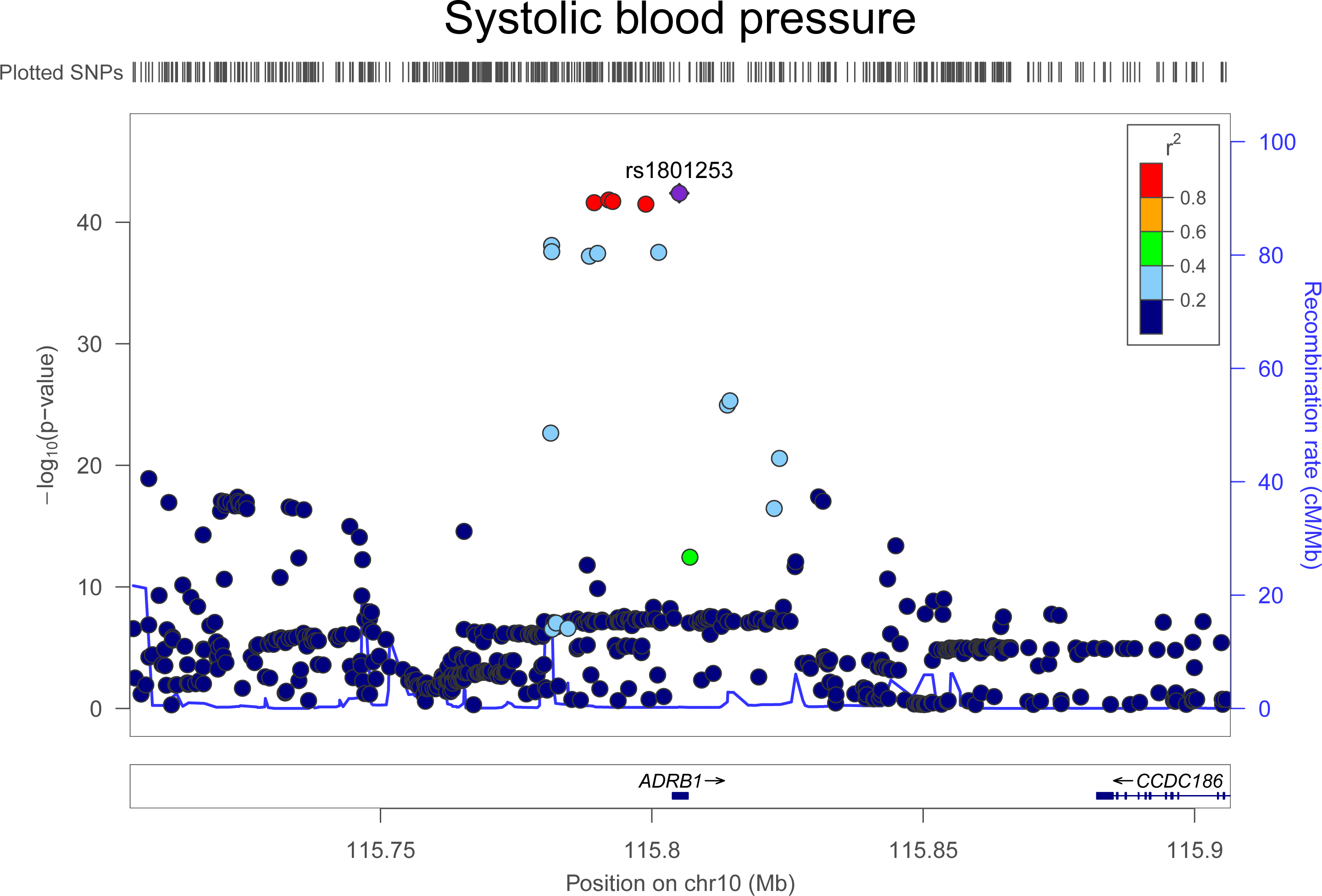

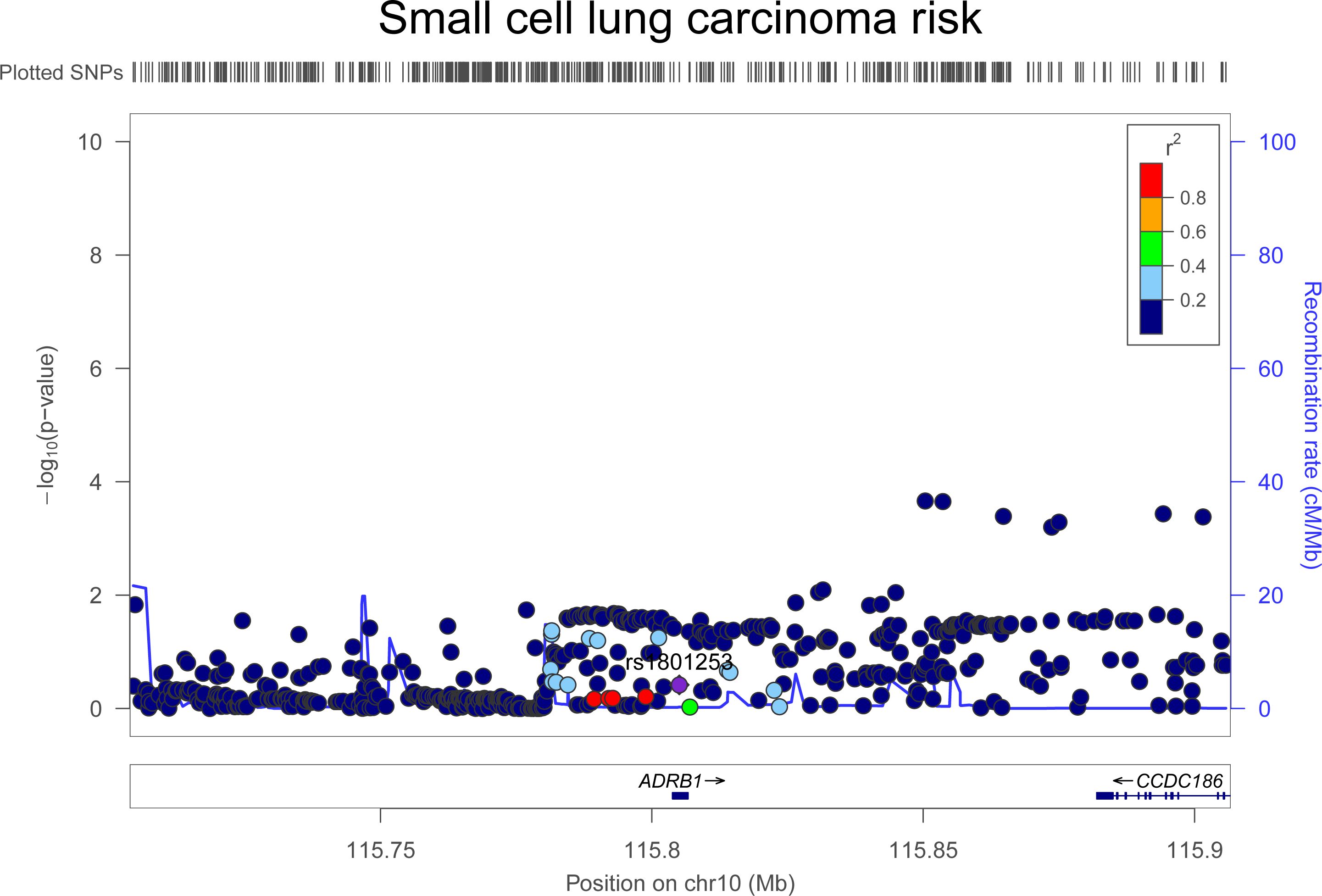

**Table 3.** Association between genetically-proxied ADRB1 inhibition and risk of overall and subtypespecific breast, colorectal, prostate, and lung cancer risk

| **Outcome** | **N (cases, controls)** | **OR (95% CI)** | **P-value** |
| --- | --- | --- | --- |
| Breast cancer | 122,977; 105,974 | 1.01 (0.99-1.04) | 0.38 |
| ER+ Breast cancer | 69,501; 105,974 | 1.01 (0.98-1.04) | 0.44 |
| ER- Breast cancer | 21,468; 105,974 | 0.98 (0.94-1.02) | 0.38 |
| Colorectal cancer | 58,221; 67,694 | 0.98 (0.96-1.01) | 0.31 |
| Colon cancer | 32,002; 64,159 | 0.99 (0.95-1.03) | 0.63 |
| Rectal cancer | 16,212; 64,159 | 1.00 (0.95-1.04) | 0.84 |
| Lung cancer | 29,863; 55,586 | 1.01 (0.96-1.07) | 0.64 |
| Lung adenocarcinoma | 11,245; 54,619 | 0.98 (0.91-1.04) | 0.48 |
| Small cell lung carcinoma | 2,791; 20,580 | 0.87 (0.79-0.96) | 0.008 |
| Squamous cell lung cancer | 7,704; 54,763 | 0.98 (0.91-1.06) | 0.67 |
| Prostate cancer | 79,148; 61,106 | 1.00 (0.96-1.03) | 0.73 |
| Advanced prostate cancer | 15,167; 58,308 | 1.00 (0.94-1.06) | 0.97 |

OR represents the exponential change in odds of cancer per genetically proxied inhibition of ADRB1 equivalent to a 1 mmHg decrease in systolic blood pressure.

### Genetically-proxied NCC inhibition and cancer risk

There was little evidence that genetically-proxied NCC inhibition was associated with overall risk of breast, colorectal, lung, or prostate cancer (**Table 4**). In oestrogen receptor-stratified breast cancer analyses, there was evidence that NCC inhibition was associated with an increased risk of ER- breast cancer (OR equivalent to 1 mmHg lower SBP: 1.20, 95% CI 1.02-1.40; *P* = 0.03). Colocalisation analysis provided little support for NCC and ER- breast cancer association sharing a causal variant within the *SLC12A3* locus (5.6% posterior probability of a shared causal variant) (**Supplementary Table 9, Figures 5-6**).

**Table 4.** Association between genetically-proxied NCC inhibition and risk of overall and subtypespecific breast, colorectal, prostate, and lung cancer risk

| **Outcome** | **N (cases, controls)** | **OR (95% CI)** | **P-value** |
| --- | --- | --- | --- |
| Breast cancer | 122,977; 105,974 | 1.08 (0.99-1.18) | 0.08 |
| ER+ Breast cancer | 69,501; 105,974 | 1.06 (0.95-1.18) | 0.28 |
| ER- Breast cancer | 21,468; 105,974 | 1.20 (1.02-1.40) | 0.03 |
| Colorectal cancer | 58,221; 67,694 | 1.09 (0.96-1.23) | 0.19 |
| Colon cancer | 32,002; 64,159 | 1.03 (0.89-1.19) | 0.69 |
| Rectal cancer | 16,212; 64,159 | 1.13 (0.94-1.36) | 0.20 |
| Lung cancer | 29,863; 55,586 | 1.09 (0.89-1.33) | 0.38 |
| Lung adenocarcinoma | 11,245; 54,619 | 1.01 (0.81-1.26) | 0.95 |
| Small cell lung carcinoma | 2,791; 20,580 | 1.12 (0.76-1.53) | 0.57 |
| Squamous cell lung cancer | 7,704; 54,763 | 1.00 (0.78-1.29) | 0.99 |
| Prostate cancer | 79,148; 61,106 | 1.08 (0.96-1.19) | 0.18 |
| Advanced prostate cancer | 15,167; 58,308 | 1.05 (0.86-1.28) | 0.63 |

OR represents the exponential change in odds of cancer per genetically proxied inhibition of NCC equivalent to a 1 mmHg decrease in systolic blood pressure.

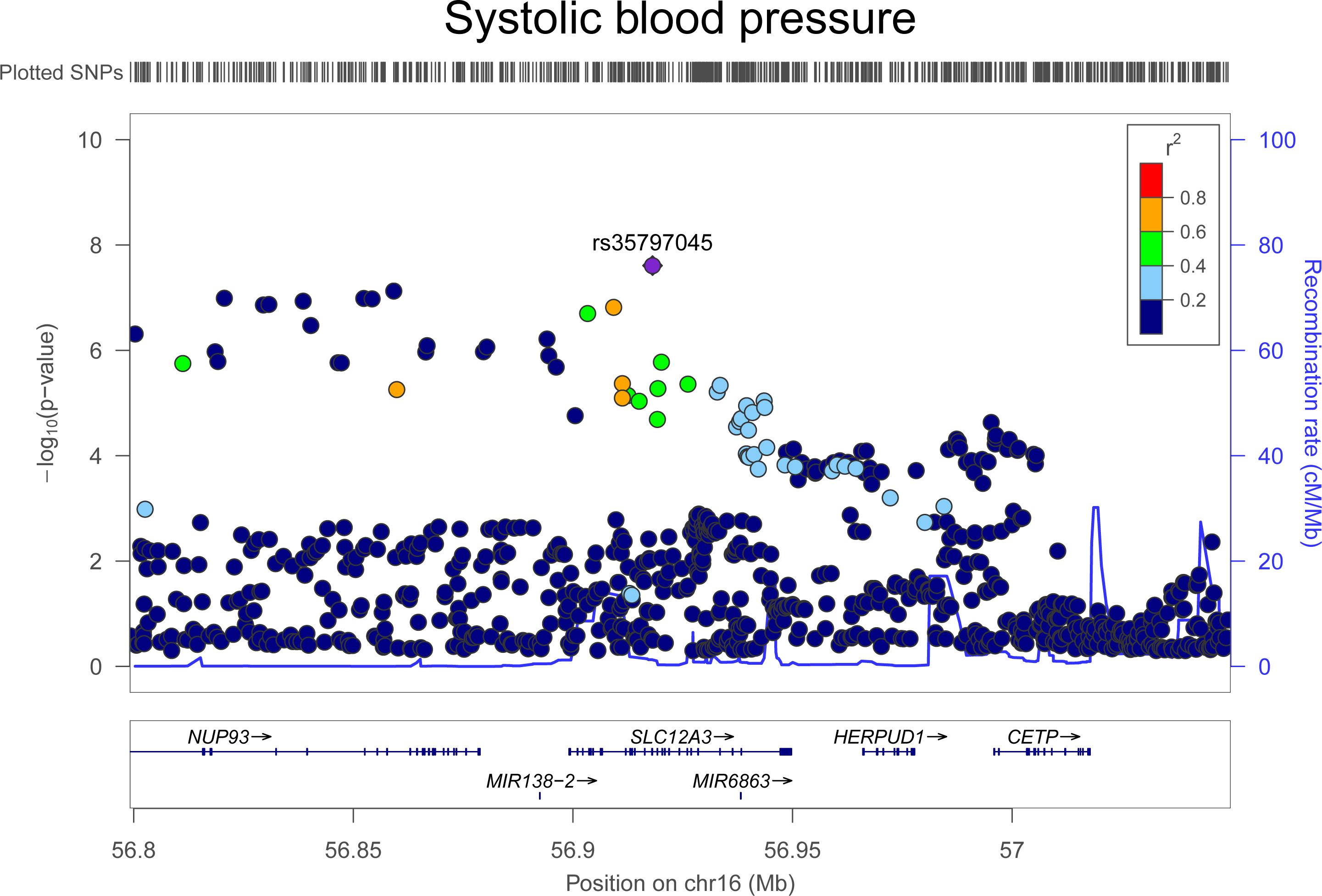

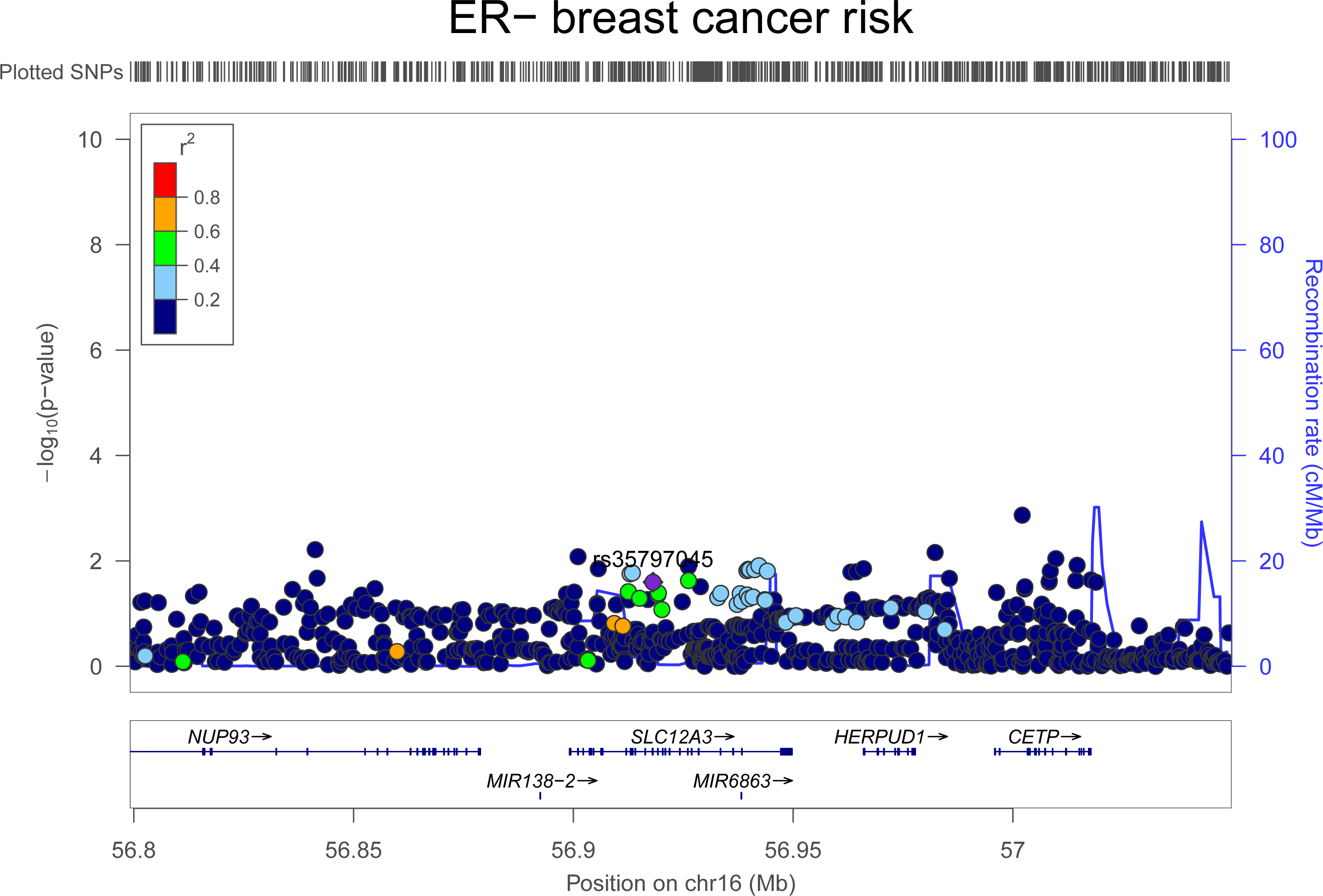

### Replication analysis in Europeans and exploratory analysis in East Asians

Findings for genetically-proxied ACE inhibition and colorectal cancer risk were replicated in an independent sample of 1,571 colorectal cancer cases and 120,006 controls of European ancestry in the Finngen consortium (1.40, 95% CI 1.02-1.92; *P* = 0.035). In analyses of 23,572 colorectal cancer cases and 48,700 controls of East Asian descent, there was little evidence of association of genetically-proxied ACE inhibition and colorectal cancer risk (OR 0.97, 95% CI 0.88-1.07; *P* = 0.59).

### Colon gene expression analysis

In transcriptome-wide analyses, genetically-proxied serum ACE inhibition was most strongly associated with *ACE* expression levels in the colon (*P* = 2.29 x 10^-31^). Genetically-proxied *ACE* expression in the colon was associated with increased odds of colorectal cancer (OR per SD increase in expression: 1.02, 95% CI 1.00-1.04; *P* = 0.01). However, colocalisation analysis suggested that colon *ACE* expression and colorectal cancer risk were unlikely to share a causal variant within the *ACE* locus (7.6% posterior probability of a shared causal variant) (**Supplementary Table 10, Figures 7-8**). Genetically-proxied serum ACE inhibition was also associated with expression levels of C*YB561* (*P* = 8.28 x 10^-11^) and *FTSJ3* (*P* = 2.95 x 10^-10^) in the colon after correction for multiple testing. *ACE*, *CYB561*, and *FTSJ3* are neighbouring genes on chromosome 17 suggesting that associations between the ACE wGRS and *CYB561* and *FTSJ3* could be driven through their co- expression. Genetically-proxied *CYB561* expression in the colon was associated with increased odds of colorectal cancer (OR per SD increase in expression: 1.06, 95% CI 1.02-1.10; *P* = 0.005). However, multivariable Mendelian randomization analysis examining the association of genetically-proxied ACE inhibition with colorectal cancer risk adjusting for *CYB561* expression in the colon was consistent with univariable analyses (OR 1.13, 95% CI: 0.96-1.32; *P* = 0.14). Genetically-proxied *FTSJ3* expression in the colon was not associated with odds of colorectal cancer (OR per SD increase in expression: 1.00, 95% CI 0.98-1.03; *P* = 0.77).

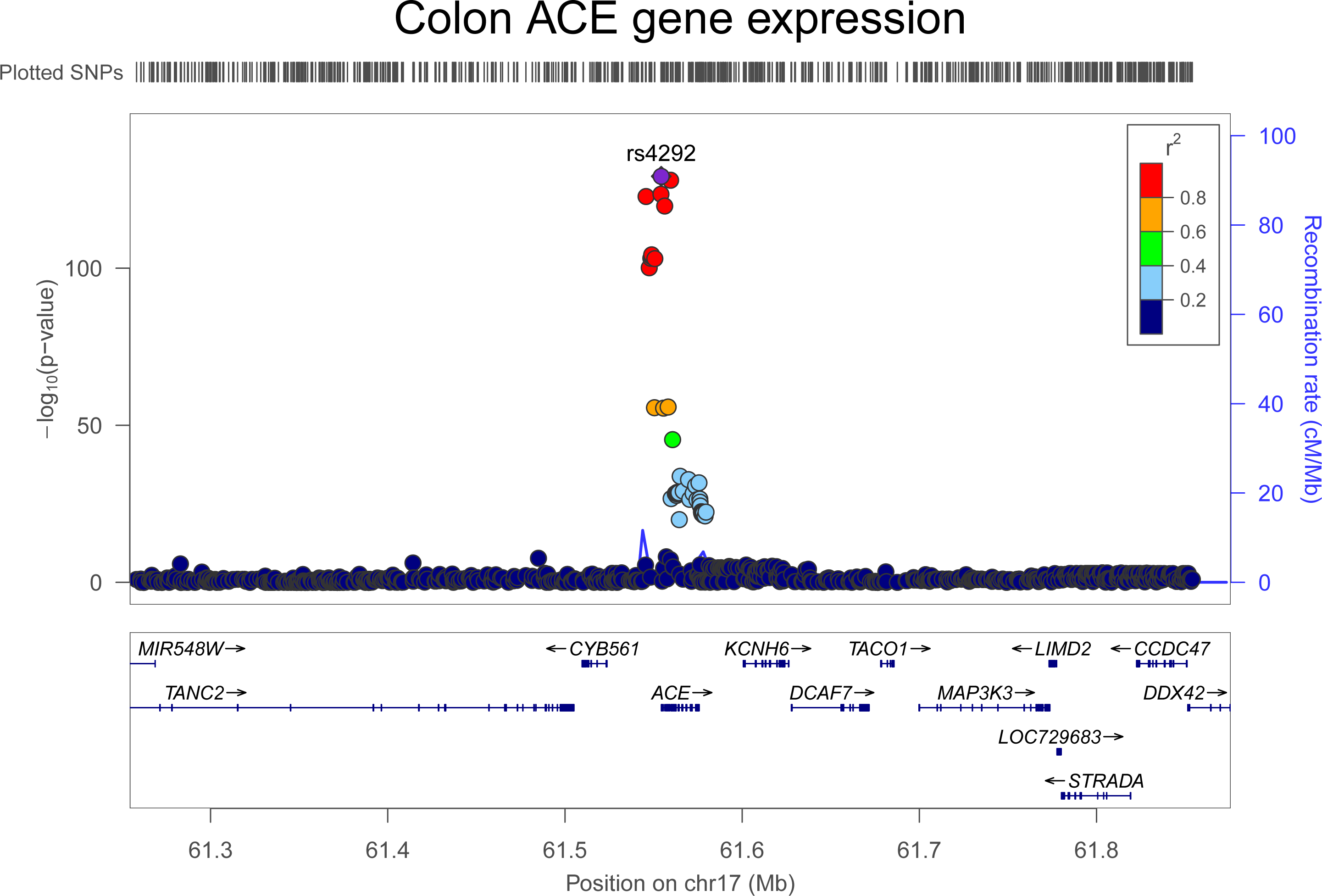

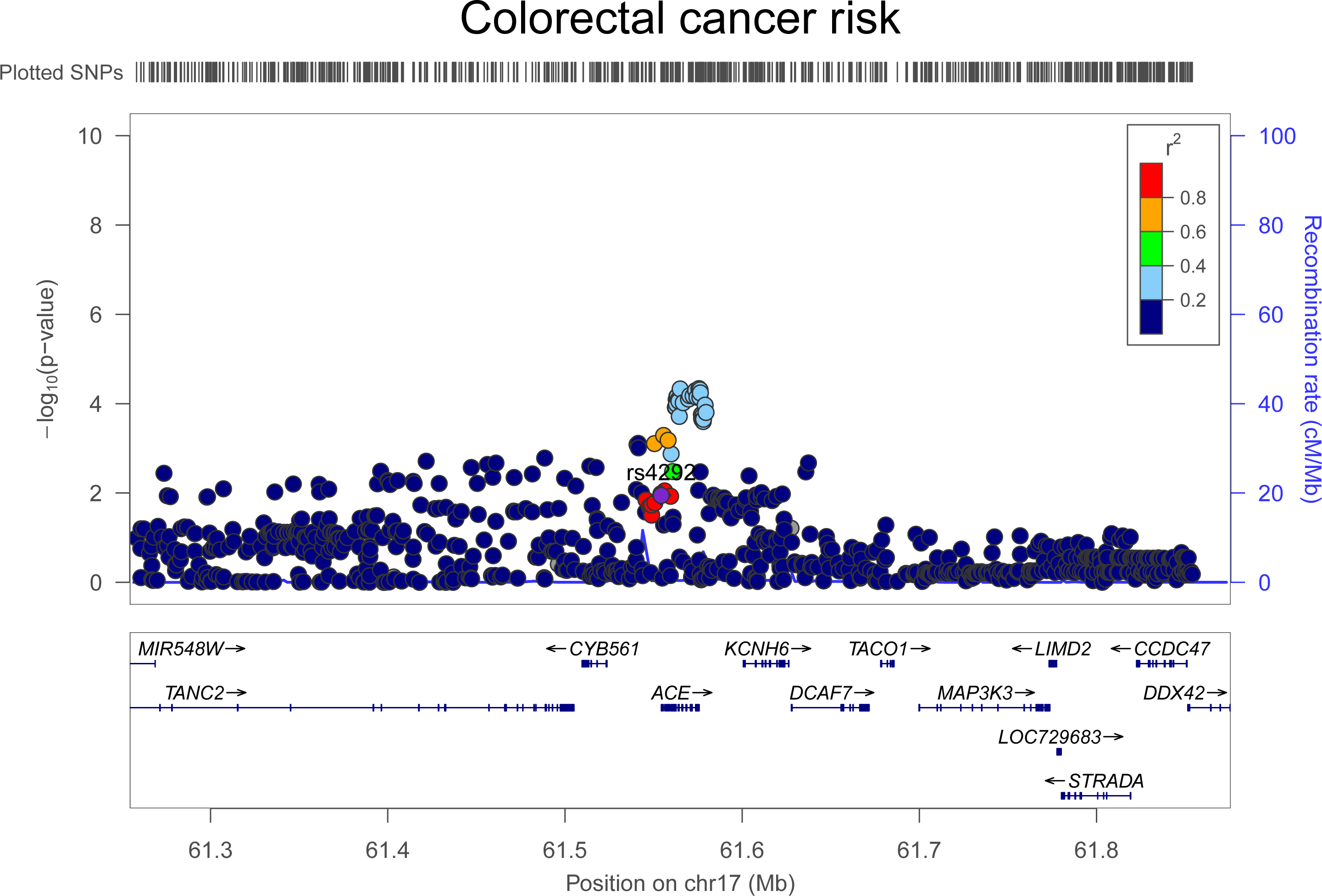

In gene-set enrichment analysis of genes whose expression was associated with genetically-proxied serum ACE inhibition (*P* < 5.0 x 10^-3^), there was evidence for enrichment of expression of genes relating to memory CD8 T cells (as compared to effector CD8 T cells) in the immunologic signatures database (GSE10239) (*P* = 1.35 x 10^-6^) but little evidence for expression of other gene-sets or pathways after correction for multiple testing.

In co-expression network analysis, 30 distinct modules were defined. *ACE* was in the black module along with another 659 genes. This module was correlated with the ACE wGRS (r= -0.11; *P* = 0.03). Gene-set enrichment analysis of genes located in the black module showed evidence of enrichment in susceptibility genes for colorectal cancer (*P* = 1.00 x 10^-3^).

Complete findings from transcriptome-wide Mendelian randomization and gene-set enrichment analyses, along with genes from the black module from co-expression network analysis are presented in **Supplementary Tables 11-14**.

## Discussion

In this Mendelian randomization analysis of up to 266,018 cancer cases and 291,224 controls, genetically-proxied long-term ACE inhibition was associated with an increased risk of colorectal cancer. This association was restricted to cancer of the colon, with similar associations across the proximal and distal colon. There was little evidence to support associations of genetically-proxied ACE inhibition with risk of other cancers. Genetically-proxied ADRB1 and NCC inhibition were not associated with risk of breast, colorectal, lung or prostate cancer.

Our findings for genetically-proxied ACE inhibition and colorectal cancer risk are not consistent with some previous conventional observational analyses. A meta-analysis of seven observational studies reported a protective association of ACE inhibitor use with colorectal cancer risk (OR 0.81 95% CI 0.70-0.92), though with substantial heterogeneity across studies (*I*^2^=71.1%)[65]. Interpretation of these findings is complicated by variable use of prevalent drug users, heterogenous comparator groups (both active controls and non-drug users), and the potential for immortal time bias across most included studies. Further, this meta-analysis did not include data from an earlier large Danish population-based case-control analysis with 15,560 colorectal cancer cases and 62,525 controls which reported an increased risk of colorectal cancer (OR 1.30, 95% CI 1.22-1.39) among long-term users of ACE inhibitors (≥ 1,000 daily doses within 5 years of study entry), as compared to never-users[7].

The potential mechanisms underpinning an association between genetically-proxied ACE inhibition and colorectal cancer risk are unclear. ACE is a multi-faceted enzyme, capable of cleaving several different peptide substrates with potential roles in carcinogenesis[66]. Along with ACE inhibition leading to an accumulation of bradykinin and substance P, both potential inducers of tumour proliferation, ACE inhibition can also lead to an increase in Ac-SDKP, an endogenous anti-fibrotic peptide which is capable of inducing angiogenesis[67]. The observed restriction of an association of genetically-proxied ACE inhibition with risk of colon, but not rectal, cancer is consistent with evidence that mRNA and protein levels of ACE are enriched in the colon but not in rectal tissue[68]. There was limited evidence of association of genetically-proxied ACE inhibition with distinct gene expression profiles in transcriptome-wide analyses. However, gene-set enrichment analysis of these findings suggested enriched expression of genes involved in immunological pathways relating to memory CD8 T cells and co-expression network analysis identified ACE expression in a cluster of co-expressed genes enriched for colorectal cancer risk susceptibility genes (e.g. *LAMA5*, *PNKD*, *TOX2*, *PLEKHG6*)[69]. These findings suggest potential future avenues of exploration to uncover mechanistic pathways linking ACE with colorectal cancer risk.

Meta-analyses of randomised trials have not reported increased rates of cancer among ACE inhibitor users, though these analyses have not reported findings separately for colorectal cancer[3, 70].

Potential discrepancies in findings for colorectal cancer between this Mendelian randomization analysis and previous clinical trials could reflect the relatively short duration of these trials (median follow-up of 3.5 years) given long induction periods of colorectal cancer. For example, the “adeno- carcinoma sequence” proposes that transformation of normal colorectal epithelium to an adenoma and ultimately to invasive and metastatic cancer may occur over the course of several decades[71, 72].

Consistent with this long induction period, in randomised controlled trials examining the chemopreventive effect of aspirin on colorectal cancer risk, protective effects of aspirin are not seen until seven years after initiation of treatment, with clear risk reductions becoming apparent only after ten years of follow-up [73]. Alternatively, it may be possible that an effect of ACE inhibition on cancer is restricted solely to the earliest stages of the adenoma-carcinoma sequence and therefore may not influence cancer risk among largely middle-aged participants of clinical trials if dysplasia is already present. Finally, it is possible that lower levels of circulating ACE concentrations may influence colorectal cancer risk only during a particular critical or sensitive period of the life-course (e.g. in childhood or adolescence), given some evidence to suggest a potential role of early-life factors in colorectal carcinogenesis[74].

Our largely null findings for genetically-proxied ACE inhibition and risk of breast, lung, and prostate cancer risk are not consistent with some previous observational reports that compared ACE inhibitor users to non-users or to users of β blockers or thiazide diuretics[7, 9, 15]. However, our findings for genetically-proxied ACE inhibition are in agreement with those from short-term randomised controlled trials for these site-specific cancers and suggest that long-term use of these drugs may not influence cancer risk, though we cannot rule out small effects from their long-term use[3]. Likewise, our findings for ADRB1 and NCC are in agreement with short-term trial data reporting no association of β blockers and thiazide diuretics use with overall cancer risk[2].

Strengths of this analysis include the use of *cis-*acting variants in genes encoding antihypertensive drug targets to proxy inhibition of these targets which should minimise confounding, the employment of various sensitivity analyses to rigorously assess for violations of Mendelian randomization assumptions, and the use of a summary-data Mendelian randomization approach which permitted us to leverage large-scale genetic data from several cancer GWAS consortia, enhancing statistical power and precision of causal estimates. As with prior Mendelian randomization analyses of antihypertensive drug targets that used similar approaches to instrument construction to our analysis, the general concordance of estimates of the effect of these instruments on cardio-metabolic endpoints with those reported in prior clinical trials for these medications supports the plausibility of these instruments[75, 76]. Finally, the use of germline genetic variants as proxies for antihypertensive drug targets facilitated evaluation of the effect of the long-term inhibition of these targets, which may be more representative of the typically decades-long use of antihypertensive therapy as compared to periods of medication use typically examined in conventional observational studies and randomised trials.

There are several limitations to these analyses. First, Mendelian randomization analyses are restricted to examining on-target (i.e. target-mediated) effects of therapeutic interventions. Second, statistical power was likely limited in some analyses of less common cancer subtypes. Limited statistical power in analyses of genetically-proxied ACE inhibition and colorectal cancer risk in East Asians (instrumented by rs4343) may also have accounted for the lack of association between these traits within this population. Third, while these analyses did not account for previously reported associations of genetically-proxied elevated systolic blood pressure with antihypertensive medication use within the colorectal cancer datasets analysed, such correction would be expected to strengthen, rather than attenuate, findings presented in this study[77]. Fourth, though mixed-ancestry GWAS were used to construct instruments for serum ACE concentrations, effect allele frequencies for variants used in this instrument were similar across European and Latin American ancestry participants, suggesting that Mendelian randomization findings were unlikely to be influenced by confounding through residual population stratification. Fifth, effect estimates presented make the additional assumptions of linearity and the absence of gene-environment and gene-gene interactions. Sixth, our genetically-proxied ADRB1 findings are of greater relevance to second generation β blockers (e.g. atenolol and metoprolol) which selectively inhibit ADRB1 as compared to first generation β blockers (e.g. propranolol and nadolol) which equally inhibit ADRB1 and ADRB2[78]. Future Mendelian randomization analyses examining the potential effects of long-term first generation β blocker use incorporating both *ADRB1* and *ADRB2* variants is warranted. Seventh, we cannot rule out findings presented being influenced by canalization (i.e. compensatory processes being generated during development that counter the phenotypic impact of genetic variants being used as instruments). Finally, while various sensitivity analyses were performed to examine exchangeability and exclusion restriction violations, these assumptions are unverifiable.

Colorectal cancer is the third most common cause of cancer globally [79]. Given the prevalence of ACE inhibitor use in high- and middle-income countries and growing use in low-income countries, and the often long-term nature of antihypertensive therapy, these findings, if replicated in subsequent clinical trials, may have important implications for choice of antihypertensive therapy[4]. Importantly, given that hypertension is more prevalent among those who are overweight or obese (risk factors for colorectal cancer), these findings suggest that long-term use of this medication could increase colorectal cancer risk among populations who are already at elevated risk of this disease. Further work is warranted to unravel molecular mechanisms underpinning the association of ACE with colorectal cancer risk. In addition, extension of the analyses presented in this study to a survival framework could inform on whether concurrent use of ACE inhibitors may have an adverse effect on prognosis among colorectal cancer patients. Finally, findings from this analysis should be “triangulated” by employing other epidemiological designs with orthogonal (i.e. non-overlapping) sources of bias to each other to further evaluate the association of ACE inhibition and colorectal cancer risk[80].

## Conclusion

Our Mendelian randomization analyses suggest that genetically-proxied long-term inhibition of ACE may increase risk of colorectal cancer. Evaluation of ACE inhibitor use in randomised controlled trials with sufficient follow-up data can inform on the long-term safety of these medications. Our findings provide human genetic support to short-term randomised trials that long-term use of β blockers and thiazide diuretics may not influence risk of common cancers.

## Data Availability

This analysis used both restricted and publicly-available summary genetic association data. Further information on applying to access restricted data from this analysis can be obtained by visiting the following websites: GECCO (https://www.fredhutch.org/en/research/divisions/public-health-sciences-division/research/cancer-prevention/genetics-epidemiology-colorectal-cancer-consortium-gecco.html), INTEGRAL-ILCCO (https://ilcco.iarc.fr/) and PRACTICAL (http://practical.icr.ac.uk/blog/).

## Acknowledgements

The authors would like to thank the participants of the individual studies contributing to the BCAC, GECCO, CORECT, CCFR, INTEGRAL-ILCCO, PRACTICAL, Finngen, BioBank Japan, Asia Colorectal Cancer Consortium, ICBP, DIAGRAM, CARDIoGRAMplusC4D, and MEGASTROKE consortia and the Genetic Epidemiology Research on Adult Health, UK Biobank, and the Korean National Cancer Center CRC Study 2. The authors would also like to acknowledge the investigators of these consortia and studies for generating the data used for this analysis. The authors would like to acknowledge the following investigators of the OncoArray and GAME-ON1KG INTEGRAL-ILCCO analyses: Maria Teresa Landi, Victoria Stevens, Ying Wang, Demetrios Albanes, Neil Caporaso, Paul Brennan, Christopher I Amos, Sanjay Shete, Rayjean J Hung, Heike Bickeböller, Angela Risch, Richard Houlston, Stephen Lam, Adonina Tardon, Chu Chen, Stig E Bojesen, Mattias Johansson, H-Erich Wichmann, David Christiani, Gadi Rennert, Susanne Arnold, John K. Field, Loic Le Marchand, Olle Melander, Hans Brunnström , Geoffrey Liu, Angeline Andrew, Lambertus A Kiemeney, Hongbing Shen, Shan Zienolddiny, Kjell Grankvist, Mikael Johansson, M Dawn Teare, Yun-Chul Hong, Jian-Min Yuan, Philip Lazarus, Matthew B Schabath, Melinda C Aldrich.

Consortia acknowledgments:

ASTERISK: We are very grateful to Dr. Bruno Buecher without whom this project would not have existed. We also thank all those who agreed to participate in this study, including the patients and the healthy control persons, as well as all the physicians, technicians and students.

CCFR: The Colon CFR graciously thanks the generous contributions of their study participants, dedication of study staff, and the financial support from the U.S. National Cancer Institute, without which this important registry would not exist.

CLUE: We appreciate the continued efforts of the staff members at the Johns Hopkins George W. Comstock Center for Public Health Research and Prevention in the conduct of the CLUE II study. We thank the participants in CLUE. Cancer incidence data for CLUE were provided by the Maryland Cancer Registry, Center for Cancer Surveillance and Control, Maryland Department of Health, 201 W.

Preston Street, Room 400, Baltimore, MD 21201, http://phpa.dhmh.maryland.gov/cancer, 410-767- 4055. We acknowledge the State of Maryland, the Maryland Cigarette Restitution Fund, and the National Program of Cancer Registries of the Centers for Disease Control and Prevention for the funds that support the collection and availability of the cancer registry data.

COLON and NQplus: the authors would like to thank the COLON and NQplus investigators at Wageningen University & Research and the involved clinicians in the participating hospitals.

CORSA: We kindly thank all individuals who agreed to participate in the CORSA study. Furthermore, we thank all cooperating physicians and students and the Biobank Graz of the Medical University of Graz.

CPS-II: The authors thank the CPS-II participants and Study Management Group for their invaluable contributions to this research. The authors would also like to acknowledge the contribution to this study from central cancer registries supported through the Centers for Disease Control and Prevention National Program of Cancer Registries, and cancer registries supported by the National Cancer Institute Surveillance Epidemiology and End Results program.

Czech Republic CCS: We are thankful to all clinicians in major hospitals in the Czech Republic, without whom the study would not be practicable. We are also sincerely grateful to all patients participating in this study.

DACHS: We thank all participants and cooperating clinicians, and Ute Handte-Daub, Utz Benscheid, Muhabbet Celik and Ursula Eilber for excellent technical assistance.

EDRN: We acknowledge all the following contributors to the development of the resource: University of Pittsburgh School of Medicine, Department of Gastroenterology, Hepatology and Nutrition: Lynda Dzubinski; University of Pittsburgh School of Medicine, Department of Pathology: Michelle Bisceglia; and University of Pittsburgh School of Medicine, Department of Biomedical Informatics.

EPIC: Where authors are identified as personnel of the International Agency for Research on Cancer/World Health Organization, the authors alone are responsible for the views expressed in this article and they do not necessarily represent the decisions, policy or views of the International Agency for Research on Cancer/World Health Organization.

EPICOLON: We are sincerely grateful to all patients participating in this study who were recruited as part of the EPICOLON project. We acknowledge the Spanish National DNA Bank, Biobank of Hospital Clínic–IDIBAPS and Biobanco Vasco for the availability of the samples. The work was carried out (in part) at the Esther Koplowitz Centre, Barcelona.

Harvard cohorts (HPFS, NHS, PHS): The study protocol was approved by the institutional review boards of the Brigham and Women’s Hospital and Harvard T.H. Chan School of Public Health, and those of participating registries as required.We acknowledge Channing Division of Network Medicine,

Department of Medicine, Brigham and Women’s Hospital as home of the NHS.We would like to thank the participants and staff of the HPFS, NHS and PHS for their valuable contributions as well as the following state cancer registries for their help: AL, AZ, AR, CA, CO, CT, DE, FL, GA, ID, IL, IN, IA, KY, LA, ME, MD, MA, MI, NE, NH, NJ, NY, NC, ND, OH, OK, OR, PA, RI, SC, TN, TX, VA, WA, WY.The authors assume full responsibility for analyses and interpretation of these data. Kentucky: We would like to acknowledge the staff at the Kentucky Cancer Registry.

LCCS: We acknowledge the contributions of Jennifer Barrett, Robin Waxman, Gillian Smith and Emma Northwood in conducting this study.

NCCCS I & II: We would like to thank the study participants, and the NC Colorectal Cancer Study staff.

NSHDS investigators thank the Västerbotten Intervention Programme, the Northern Sweden MONICA study, the Biobank Research Unit at Umeå University and Biobanken Norr at Region Västerbotten for providing data and samples and acknowledge the contribution from Biobank Sweden, supported by the Swedish Research Council.

PLCO: The authors thank the PLCO Cancer Screening Trial screening center investigators and the staff from Information Management Services Inc and Westat Inc. Most importantly, we thank the study participants for their contributions that made this study possible. Cancer incidence data have been provided by the District of Columbia Cancer Registry, Georgia Cancer Registry, Hawaii Cancer Registry, Minnesota Cancer Surveillance System, Missouri Cancer Registry, Nevada Central Cancer Registry, Pennsylvania Cancer Registry, Texas Cancer Registry, Virginia Cancer Registry, and Wisconsin Cancer Reporting System. All are supported in part by funds from the Center for Disease Control and Prevention, National Program for Central Registries, local states or by the National Cancer Institute, Surveillance, Epidemiology, and End Results program. The results reported here and the conclusions derived are the sole responsibility of the authors.

SCCFR: The authors would like to thank the study participants and staff of the Seattle Colon Cancer Family Registry and the Hormones and Colon Cancer study (CORE Studies).

SEARCH: We thank the SEARCH team

SELECT: We thank the research and clinical staff at the sites that participated on SELECT study, without whom the trial would not have been successful. We are also grateful to the 35,533 dedicated men who participated in SELECT.

WHI: The authors thank the WHI investigators and staff for their dedication, and the study participants for making the program possible. A full listing of WHI investigators can be found at: http://www.whi.org/researchers/Documents%20%20Write%20a%20Paper/WHI%20Investigator%20Short%20List.pdf

## Funding

JY is supported by a Cancer Research UK Population Research Postdoctoral Fellowship (C68933/A28534). JY, EEV, VT, GDS, and RMM are supported by Cancer Research UK (C18281/A29019) programme grant (the Integrative Cancer Epidemiology Programme). JY, TGR, VMW, EEV, VT, GDS, and RMM are part of the Medical Research Council Integrative Epidemiology Unit at the University of Bristol which is supported by the Medical Research Council (MC_UU_00011/1, MC_UU_00011/3, MC_UU_00011/6, and MC_UU_00011/4) and the University of Bristol. RMM is also supported by the NIHR Bristol Biomedical Research Centre which is funded by the NIHR and is a partnership between University Hospitals Bristol NHS Foundation Trust and the University of Bristol. Department of Health and Social Care disclaimer: The views expressed are those of the author(s) and not necessarily those of the NHS, the NIHR or the Department of Health and Social Care. EEV is supported by Diabetes UK (17/0005587) and the World Cancer Research Fund (WCRF UK), as part of the World Cancer Research Fund International grant programme (IIG_2019_2009). MOS received a post-doctoral fellowship from the Spanish Association Against Cancer (AECC) Scientific Foundation. VDO, MOS, and VM are part of the group 55 of CIBERESP and AGAUR 2017SGR723. CIA is a Cancer Prevention Research Institute of Texas Research Scholar and is supported by RR170048. INTEGRAL-ILCCO is supported by a National Institutes of Health grant (U19 CA203654). The International Lung Cancer Consortium is supported by a National Cancer Institute grant (U19CA203654). GC is supported by R01 NIH/NCI CA143237 and CA204279.

Consortia funding:

The breast cancer genome-wide association analyses were supported by the Government of Canada through Genome Canada and the Canadian Institutes of Health Research, the ‘Ministère de l’Économie, de la Science et de l’Innovation du Québec’ through Genome Québec and grant PSR-SIIRI-701, The National Institutes of Health (U19 CA148065, X01HG007492), Cancer Research UK (C1287/A10118, C1287/A16563, C1287/A10710) and The European Union (HEALTH-F2-2009-223175 and H2020 633784 and 634935). All studies and funders are listed in Michailidou et al (Nature, 2017).

Genetics and Epidemiology of Colorectal Cancer Consortium (GECCO): National Cancer Institute, National Institutes of Health, U.S. Department of Health and Human Services (U01 CA137088, R01 CA059045, R01 CA201407, R01 CA244588). This research was funded in part through the NIH/NCI Cancer Center Support Grant P30 CA015704. Scientific Computing Infrastructure at Fred Hutch funded by ORIP grant S10OD028685.

The ATBC Study is supported by the Intramural Research Program of the U.S. National Cancer Institute, National Institutes of Health, Department of Health and Human Services.

CLUE funding was from the National Cancer Institute (U01 CA86308, Early Detection Research Network; P30 CA006973), National Institute on Aging (U01 AG18033), and the American Institute for Cancer Research. The content of this publication does not necessarily reflect the views or policies of the Department of Health and Human Services, nor does mention of trade names, commercial products, or organizations imply endorsement by the US government.

ColoCare: This work was supported by the National Institutes of Health (grant numbers R01 CA189184 (Li/Ulrich), U01 CA206110 (Ulrich/Li/Siegel/Figueiredo/Colditz, 2P30CA015704- 40 (Gilliland), R01 CA207371 (Ulrich/Li)), the Matthias Lackas-Foundation, the German Consortium for Translational Cancer Research, and the EU TRANSCAN initiative.

The Colon Cancer Family Registry (CCFR, www.coloncfr.org) is supported in part by funding from the National Cancer Institute (NCI), National Institutes of Health (NIH) (award U01 CA167551). The CCFR Set-1 (Illumina 1M/1M-Duo) and Set-2 (Illumina Omni1-Quad) scans were supported by NIH awards U01 CA122839 and R01 CA143247 (to GC). The CCFR Set-3 (Affymetrix Axiom CORECT Set array) was supported by NIH award U19 CA148107 and R01 CA81488 (to SBG). The CCFR Set- 4 (Illumina OncoArray 600K SNP array) was supported by NIH award U19 CA148107 (to SBG) and by the Center for Inherited Disease Research (CIDR), which is funded by the NIH to the Johns Hopkins University, contract number HHSN268201200008I. The content of this manuscript does not necessarily reflect the views or policies of the NCI, NIH or any of the collaborating centers in the Colon Cancer Family Registry (CCFR), nor does mention of trade names, commercial products, or organizations imply endorsement by the US Government, any cancer registry, or the CCFR.

COLON: The COLON study is sponsored by Wereld Kanker Onderzoek Fonds, including funds from grant 2014/1179 as part of the World Cancer Research Fund International Regular Grant Programme, by Alpe d’Huzes and the Dutch Cancer Society (UM 2012–5653, UW 2013-5927, UW2015-7946), and by TRANSCAN (JTC2012-MetaboCCC, JTC2013-FOCUS). The Nqplus study is sponsored by a ZonMW investment grant (98-10030); by PREVIEW, the project PREVention of diabetes through lifestyle intervention and population studies in Europe and around the World (PREVIEW) project which received funding from the European Union Seventh Framework Programme (FP7/2007–2013) under grant no. 312057; by funds from TI Food and Nutrition (cardiovascular health theme), a public– private partnership on precompetitive research in food and nutrition; and by FOODBALL, the Food Biomarker Alliance, a project from JPI Healthy Diet for a Healthy Life.

Colorectal Cancer Transdisciplinary (CORECT) Study: The CORECT Study was supported by the National Cancer Institute, National Institutes of Health (NCI/NIH), U.S. Department of Health and Human Services (grant numbers U19 CA148107, R01 CA81488, P30 CA014089, R01 CA197350; P01 CA196569; R01 CA201407) and National Institutes of Environmental Health Sciences, National Institutes of Health (grant number T32 ES013678).

CORSA: The CORSA study was funded by Austrian Research Funding Agency (FFG) BRIDGE (grant 829675, to Andrea Gsur), the “Herzfelder’sche Familienstiftung” (grant to Andrea Gsur) and was supported by COST Action BM1206.

CPS-II: The American Cancer Society funds the creation, maintenance, and updating of the Cancer Prevention Study-II (CPS-II) cohort. This study was conducted with Institutional Review Board approval.

CRCGEN: Colorectal Cancer Genetics & Genomics, Spanish study was supported by Instituto de Salud Carlos III, co-funded by FEDER funds –a way to build Europe– (grants PI14-613 and PI09- 1286), Agency for Management of University and Research Grants (AGAUR) of the Catalan

Government (grant 2017SGR723), and Junta de Castilla y León (grant LE22A10-2). Sample collection of this work was supported by the Xarxa de Bancs de Tumors de Catalunya sponsored by Pla Director d’Oncología de Catalunya (XBTC), Plataforma Biobancos PT13/0010/0013 and ICOBIOBANC, sponsored by the Catalan Institute of Oncology.

Czech Republic CCS: This work was supported by the Grant Agency of the Czech Republic (18- 09709S, 20-03997S), by the Grant Agency of the Ministry of Health of the Czech Republic (grants AZV NV18/03/00199 and AZV NV19-09-00237), and Charles University grants Unce/Med/006 and Progress Q28/LF1.

DACHS: This work was supported by the German Research Council (BR 1704/6-1, BR 1704/6-3, BR 1704/6-4, CH 117/1-1, HO 5117/2-1, HE 5998/2-1, KL 2354/3-1, RO 2270/8-1 and BR 1704/17-1), the Interdisciplinary Research Program of the National Center for Tumor Diseases (NCT), Germany, and the German Federal Ministry of Education and Research (01KH0404, 01ER0814, 01ER0815, 01ER1505A and 01ER1505B).

DALS: National Institutes of Health (R01 CA48998 to M. L. Slattery).

EDRN: This work is funded and supported by the NCI, EDRN Grant (U01 CA 84968-06).

EPIC: The coordination of EPIC is financially supported by International Agency for Research on Cancer (IARC) and also by the Department of Epidemiology and Biostatistics, School of Public Health, Imperial College London which has additional infrastructure support provided by the NIHR Imperial Biomedical Research Centre (BRC). The national cohorts are supported by: Danish Cancer Society (Denmark); Ligue Contre le Cancer, Institut Gustave Roussy, Mutuelle Générale de l’Education Nationale, Institut National de la Santé et de la Recherche Médicale (INSERM) (France); German Cancer Aid, German Cancer Research Center (DKFZ), German Institute of Human Nutrition Potsdam- Rehbruecke (DIfE), Federal Ministry of Education and Research (BMBF) (Germany); Associazione Italiana per la Ricerca sul Cancro-AIRC-Italy, Compagnia di SanPaolo and National Research Council (Italy); Dutch Ministry of Public Health, Welfare and Sports (VWS), Netherlands Cancer Registry (NKR), LK Research Funds, Dutch Prevention Funds, Dutch ZON (Zorg Onderzoek Nederland), World Cancer Research Fund (WCRF), Statistics Netherlands (The Netherlands); Health Research Fund (FIS) - Instituto de Salud Carlos III (ISCIII), Regional Governments of Andalucía, Asturias, Basque Country, Murcia and Navarra, and the Catalan Institute of Oncology - ICO (Spain); Swedish Cancer Society, Swedish Research Council and County Councils of Skåne and Västerbotten (Sweden); Cancer Research UK (14136 to EPIC-Norfolk; C8221/A29017 to EPIC-Oxford), Medical Research Council (1000143 to EPIC-Norfolk; MR/M012190/1 to EPIC-Oxford). (United Kingdom).

EPICOLON: This work was supported by grants from Fondo de Investigación Sanitaria/FEDER (PI08/0024, PI08/1276, PS09/02368, P111/00219, PI11/00681, PI14/00173, PI14/00230, PI17/00509, 17/00878, PI20/00113, PI20/00226, Acción Transversal de Cáncer), Xunta de Galicia (PGIDIT07PXIB9101209PR), Ministerio de Economia y Competitividad (SAF07-64873, SAF 2010- 19273, SAF2014-54453R), Fundación Científica de la Asociación Española contra el Cáncer (GCB13131592CAST), Beca Grupo de Trabajo “Oncología” AEG (Asociación Española de Gastroenterología), Fundación Privada Olga Torres, FP7 CHIBCHA Consortium, Agència de Gestió d’Ajuts Universitaris i de Recerca (AGAUR, Generalitat de Catalunya, 2014SGR135, 2014SGR255, 2017SGR21, 2017SGR653), Catalan Tumour Bank Network (Pla Director d’Oncologia, Generalitat de Catalunya), PERIS (SLT002/16/00398, Generalitat de Catalunya), CERCA Programme (Generalitat de Catalunya) and COST Action BM1206 and CA17118. CIBERehd is funded by the Instituto de Salud Carlos III.

ESTHER/VERDI. This work was supported by grants from the Baden-Württemberg Ministry of Science, Research and Arts and the German Cancer Aid.

Harvard cohorts (HPFS, NHS, PHS): HPFS is supported by the National Institutes of Health (P01 CA055075, UM1 CA167552, U01 CA167552, R01 CA137178, R01 CA151993, and R35 CA197735),

NHS by the National Institutes of Health (R01 CA137178, P01 CA087969, UM1 CA186107, R01 CA151993, and R35 CA197735) and PHS by the National Institutes of Health (R01 CA042182).

Hawaii Adenoma Study: NCI grants R01 CA72520.

HCES-CRC: the Hwasun Cancer Epidemiology Study–Colon and Rectum Cancer (HCES-CRC; grants from Chonnam National University Hwasun Hospital, HCRI15011-1).

Kentucky: This work was supported by the following grant support: Clinical Investigator Award from Damon Runyon Cancer Research Foundation (CI-8); NCI R01CA136726.

LCCS: The Leeds Colorectal Cancer Study was funded by the Food Standards Agency and Cancer Research UK Programme Award (C588/A19167).

MCCS cohort recruitment was funded by VicHealth and Cancer Council Victoria. The MCCS was further supported by Australian NHMRC grants 509348, 209057, 251553 and 504711 and by infrastructure provided by Cancer Council Victoria. Cases and their vital status were ascertained through the Victorian Cancer Registry (VCR) and the Australian Institute of Health and Welfare (AIHW), including the National Death Index and the Australian Cancer Database.

MEC: National Institutes of Health (R37 CA54281, P01 CA033619, and R01 CA063464).

MECC: This work was supported by the National Institutes of Health, U.S. Department of Health and Human Services (R01 CA81488, R01 CA197350).

MSKCC: The work at Sloan Kettering in New York was supported by the Robert and Kate Niehaus Center for Inherited Cancer Genomics and the Romeo Milio Foundation. Moffitt: This work was supported by funding from the National Institutes of Health (grant numbers R01 CA189184, P30 CA076292), Florida Department of Health Bankhead-Coley Grant 09BN-13, and the University of South Florida Oehler Foundation. Moffitt contributions were supported in part by the Total Cancer Care Initiative, Collaborative Data Services Core, and Tissue Core at the H. Lee Moffitt Cancer Center & Research Institute, a National Cancer Institute-designated Comprehensive Cancer Center (grant number P30 CA076292).

NCCCS I & II: We acknowledge funding support for this project from the National Institutes of Health, R01 CA66635 and P30 DK034987.

NFCCR: This work was supported by an Interdisciplinary Health Research Team award from the Canadian Institutes of Health Research (CRT 43821); the National Institutes of Health, U.S. Department of Health and Human Serivces (U01 CA74783); and National Cancer Institute of Canada grants (18223 and 18226). The authors wish to acknowledge the contribution of Alexandre Belisle and the genotyping team of the McGill University and Génome Québec Innovation Centre, Montréal, Canada, for genotyping the Sequenom panel in the NFCCR samples. Funding was provided to Michael O. Woods by the Canadian Cancer Society Research Institute.

NSHDS: The research was supported by Biobank Sweden through funding from the Swedish Research Council (VR 2017-00650, VR 2017-01737), the Swedish Cancer Society (CAN 2017/581), Region Västerbotten (VLL-841671, VLL-833291), Knut and Alice Wallenberg Foundation (VLL- 765961), and the Lion’s Cancer Research Foundation (several grants) and Insamlingsstiftelsen, both at Umeå University.

OFCCR: The Ontario Familial Colorectal Cancer Registry was supported in part by the National Cancer Institute (NCI) of the National Institutes of Health (NIH) under award U01 CA167551 and award U01/U24 CA074783 (to SG). Additional funding for the OFCCR and ARCTIC testing and genetic analysis was through and a Canadian Cancer Society CaRE (Cancer Risk Evaluation) program grant and Ontario Research Fund award GL201-043 (to BWZ), through the Canadian Institutes of Health Research award 112746 (to TJH), and through generous support from the Ontario Ministry of Research and Innovation.

OSUMC: OCCPI funding was provided by Pelotonia and HNPCC funding was provided by the NCI (CA16058 and CA67941).

PLCO: Intramural Research Program of the Division of Cancer Epidemiology and Genetics and supported by contracts from the Division of Cancer Prevention, National Cancer Institute, NIH, DHHS. Funding was provided by National Institutes of Health (NIH), Genes, Environment and Health Initiative (GEI) Z01 CP 010200, NIH U01 HG004446, and NIH GEI U01 HG 004438.

SCCFR: The Seattle Colon Cancer Family Registry was supported in part by the National Cancer Institute (NCI) of the National Institutes of Health (NIH) under awards U01 CA167551, U01 CA074794 (to JDP), and awards U24 CA074794 and R01 CA076366 (to PAN).

SEARCH: The University of Cambridge has received salary support in respect of PDPP from the NHS in the East of England through the Clinical Academic Reserve. Cancer Research UK (C490/A16561); the UK National Institute for Health Research Biomedical Research Centres at the University of Cambridge.

SELECT: Research reported in this publication was supported in part by the National Cancer Institute of the National Institutes of Health under Award Numbers U10 CA37429 (CD Blanke), and UM1 CA182883 (CM Tangen/IM Thompson). The content is solely the responsibility of the authors and does not necessarily represent the official views of the National Institutes of Health.

SMS and REACH: This work was supported by the National Cancer Institute (grant P01 CA074184 to J.D.P. and P.A.N., grants R01 CA097325, R03 CA153323, and K05 CA152715 to P.A.N., and the

National Center for Advancing Translational Sciences at the National Institutes of Health (grant KL2 TR000421 to A.N.B.-H.)

The Swedish Low-risk Colorectal Cancer Study: The study was supported by grants from the Swedish research council; K2015-55X-22674-01-4, K2008-55X-20157-03-3, K2006-72X-20157-01-2 and the

Stockholm County Council (ALF project).

Swedish Mammography Cohort and Cohort of Swedish Men: This work is supported by the Swedish Research Council /Infrastructure grant, the Swedish Cancer Foundation, and the Karolinska Institutés Distinguished Professor Award to Alicja Wolk.

UK Biobank: This research has been conducted using the UK Biobank Resource under Application Number 8614

VITAL: National Institutes of Health (K05 CA154337).

WHI: The WHI program is funded by the National Heart, Lung, and Blood Institute, National Institutes of Health, U.S. Department of Health and Human Services through contracts HHSN268201100046C, HHSN268201100001C, HHSN268201100002C, HHSN268201100003C, HHSN268201100004C, and HHSN271201100004C.

CRUK and PRACTICAL consortium

This work was supported by the Canadian Institutes of Health Research, European Commission’s Seventh Framework Programme grant agreement n° 223175 (HEALTH-F2-2009-223175), Cancer Research UK Grants C5047/A7357, C1287/A10118, C1287/A16563, C5047/A3354, C5047/A10692,

C16913/A6135, and The National Institute of Health (NIH) Cancer Post-Cancer GWAS initiative grant: No. 1 U19 CA 148537-01 (the GAME-ON initiative).

We would also like to thank the following for funding support: The Institute of Cancer Research and The Everyman Campaign, The Prostate Cancer Research Foundation, Prostate Research Campaign UK (now PCUK), The Orchid Cancer Appeal, Rosetrees Trust, The National Cancer Research Network UK, The National Cancer Research Institute (NCRI) UK. We are grateful for support of NIHR funding to the NIHR Biomedical Research Centre at The Institute of Cancer Research and The Royal Marsden NHS Foundation Trust.

The Prostate Cancer Program of Cancer Council Victoria also acknowledge grant support from The National Health and Medical Research Council, Australia (126402, 209057, 251533, 396414, 450104, 504700, 504702, 504715, 623204, 940394, 614296,), VicHealth, Cancer Council Victoria, The Prostate Cancer Foundation of Australia, The Whitten Foundation, PricewaterhouseCoopers, and Tattersall’s. EAO, DMK, and EMK acknowledge the Intramural Program of the National Human Genome Research Institute for their support.

Genotyping of the OncoArray was funded by the US National Institutes of Health (NIH) [U19 CA 148537 for ELucidating Loci Involved in Prostate cancer SuscEptibility (ELLIPSE) project and X01HG007492 to the Center for Inherited Disease Research (CIDR) under contract number

HHSN268201200008I]. Additional analytic support was provided by NIH NCI U01 CA188392 (PI: Schumacher).

Funding for the iCOGS infrastructure came from: the European Community’s Seventh Framework Programme under grant agreement n° 223175 (HEALTH-F2-2009-223175) (COGS), Cancer Research UK (C1287/A10118, C1287/A 10710, C12292/A11174, C1281/A12014, C5047/A8384,

C5047/A15007, C5047/A10692, C8197/A16565), the National Institutes of Health (CA128978) and Post-Cancer GWAS initiative (1U19 CA148537, 1U19 CA148065 and 1U19 CA148112 - the GAME-ON initiative), the Department of Defence (W81XWH-10-1-0341), the Canadian Institutes of Health Research (CIHR) for the CIHR Team in Familial Risks of Breast Cancer, Komen Foundation for the Cure, the Breast Cancer Research Foundation, and the Ovarian Cancer Research Fund.

BPC3

The BPC3 was supported by the U.S. National Institutes of Health, National Cancer Institute (cooperative agreements U01-CA98233 to D.J.H., U01-CA98710 to S.M.G., U01-CA98216 to E.R., and U01-CA98758 to B.E.H., and Intramural Research Program of NIH/National Cancer Institute, Division of Cancer Epidemiology and Genetics).

CAPS

CAPS GWAS study was supported by the Cancer Risk Prediction Center (CRisP; www.crispcenter.org), a Linneus Centre (Contract ID 70867902) financed by the Swedish Research Council, (grant no K2010-70X-20430-04-3), the Swedish Cancer Foundation (grant no 09-0677), the Hedlund Foundation, the Soederberg Foundation, the Enqvist Foundation, ALF funds from the Stockholm County Council. Stiftelsen Johanna Hagstrand och Sigfrid Linner’s Minne, Karlsson’s Fund for urological and surgical research.

PEGASUS

PEGASUS was supported by the Intramural Research Program, Division of Cancer Epidemiology and Genetics, National Cancer Institute, National Institutes of Health.

The MEGASTROKE project received funding from sources specified at http://www.megastroke.org/acknowledgments.html

