## Supplementary Table 1 for "Genetically-proxied therapeutic inhibition of antihypertensive drug targets and risk of common cancers"

**Supplementary Methods**

**Colocalization analysis**

Colocalization tests the probability of shared causal variants between two (or more) traits. The presence of shared causal variants– as opposed to distinct causal variants that are in linkage disequilibrium with each other - is necessary in order to infer potential causality between these traits (though it is not sufficient as it does not account for horizontal pleiotropy and cannot inform on direction of association between traits). In order to examine whether there was evidence of colocalization for Mendelian randomization analyses showing nominal evidence of association (*P* < 0.05), we used the coloc package to quantify the probability of shared causal variants across SNP-drug target and SNP-cancer endpoint analyses. This package uses approximate Bayes factor (ABF) computation to generates posterior probabilities that associations between two traits represent each of the following configurations: (i) neither trait has a genetic association in the region (H_0_), (ii) only the first trait has a genetic association in the region (H_1_), (iii) only the second trait has a genetic association in the region (H_2_), (iv) both traits are associated but have different causal variants (H_3_) and (v) both traits are associated and share a single causal variant (H_4_). Colocalisation analysis was performed by generating ± 300 kb windows from the top SNP used to instrument each drug target. As a convention, a posterior probability of ≥ 0.80 was used to indicate support for a configuration tested.

An assumption of coloc is that there is at most one causal variant per trait within the genomic region examined. To evaluate this assumption we generated regional plots for SNP‐drug target and SNP–cancer associations to visually inspect whether there was evidence of single or multiple independent causal variants within the region examined.

**BarcUVa-Seq study**

Individuals included in the BarcUVa-Seq study were participants of a Spanish colorectal cancer risk screening programme who obtained a normal colonoscopy result (i.e. macroscopically normal colon tissue, with no malignant lesions). RNA-Seq was performed using biopsy data collected from the superficial epithelial mucosa at colonoscopy and DNA genotyping was performed from leukocytes of peripheral blood samples. Biopsies were obtained from the ascending (31%), transverse (32%), and descending (37%) colon. Briefly, quality control of the sequencing reads was performed using FastQC, read alignment was performed using STAR, and gene expression quantification was performed using RSEM[1-3]. For downstream analyses, TMMs were computed from counts using only genes with more than 6 counts in at least 10 samples. Genotype imputation of the SNP array was performed with Minimac 4 using the TOPMed reference panel[4-6]. We only kept SNPs with high imputation quality (R^2^ > 0.7). RNA-Seq sequencing, quality control and analysis were performed as described elsewhere[7] .

**Co-expression network and gene-set enrichment analyses**

Gene co-expression network analysis was conducted with the WGCNA R package[8]. To calculate the adjacency matrix, we selected the soft-thresholding power of 8 to approximate scale-free topology. We transformed the adjacency into a topological overlap matrix and performed hierarchical clustering of genes and dynamic tree cut before assigning genes to modules. To decrease the number of genes per module, we did not merge modules whose expression profiles were very similar. We performed correlation analysis between the summary profile (eigengene) of each module and the ACE wGRS.

Genes associated with the ACE wGRS at *P* < 5.0 x 10^-3^, as well as genes located in the black module of the co-expression network, were further examined in gene-set enrichment analyses using FUMA gene2func. This tool implements a hypergeometric test of enrichment in a wide array of curated gene-sets from various sources, such as the Molecular Signatures Database (MsigDB)[9].

Supplementary Table 1. Characteristics of systolic blood pressure lowering genetic variants in *ADRB1* in a GWAS unadjusted for body mass index

| **SNP** | **Effect Allele/Non-Effect Allele** | **Effect Allele Frequency** | **Effect (SE)** | ***P*-value** |
| --- | --- | --- | --- | --- |
| rs2782980 | T/C | 0.28 | -0.35 (0.05) | 1.27 x 10^-13^ |
| rs180940 | A/G | 0.33 | -0.30 (0.04) | 4.39 x 10^-11^ |

SNP = Single-Nucleotide Polymorphism. No SNPs at genome-wide significance (*P* < 5 x 10^-8^) were available to instrument NCC. Effect (SE) represents change in systolic blood pressure per additional copy of the effect allele.

Supplementary Table 2. Comparison of effect allele frequency for variants included as ACE instruments across European and Latin American participants in serum ACE concentrations genome-wide association study and colorectal cancer risk in participants of European ancestry

| **SNP** | **Effect Allele** | **Serum ACE (EUR)** | **Serum ACE**  **(LA)** | **Colorectal cancer (EUR)** |
| --- | --- | --- | --- | --- |
| rs4343 | A | 0.45 | 0.46 | 0.47 |
| rs12452187 | G | 0.40 | 0.39 | 0.38 |
| rs79480822 | T | 0.07 | 0.03 | 0.06 |
| rs3730025 | G | 0.01 | 0.01 | 0.01 |
| rs4365 | A | 0.03 | 0.02 | 0.04 |
| rs80311894 | G | 0.03 | 0.03 | 0.05 |
| rs4968780 | C | 0.05 | 0.06 | 0.04 |
| rs11655956 | C | 0.08 | 0.08 | 0.09 |
| rs118121655 | A | 0.04 | 0.03 | 0.03 |
| rs28656895 | T | 0.23 | 0.24 | 0.23 |
| rs11650201 | G | 0.16 | 0.18 | 0.18 |
| rs12709437 | T | 0.03 | 0.02 | 0.02 |

SNP = Single-Nucleotide Polymorphism, ACE = Angiotensin-converting enzyme, EUR = European participants, LA = Latin American participants.

Supplementary Table 3. Instrument strength estimates for drug target instruments

| **Target** | **N of SNPs** | **R^2^** | **F-stats** |
| --- | --- | --- | --- |
| ACE | 3-14 | 0.34-0.39 | 2,156.5-2,594.9 |
| ADRB1 | 2-8 | 0.00031-0.00067 | 269.1-572.2 |
| NCC | 1 | 0.0021 | 1659.9 |

Range represents r^2^ and F-stats across instruments applying a linkage disequilibrium threshold of < 0.01 to < 0.10.

Supplementary Table 4. Posterior probabilities under differing hypotheses relating the associations between serum ACE concentrations and colorectal cancer risk

| **Configuration** | **H_0_** | **H_1_** | **H_2_** | **H_3_** | **H_4_** |
| --- | --- | --- | --- | --- | --- |
|  | 1.27 x 10^-230^ | 6.16 x 10^-2^ | 5.12 x 10^-231^ | 2.40 x 10^-2^ | 0.91 |

H_0_ = neither serum ACE concentrations nor colorectal cancer risk has a genetic association in the region, H_1_ = only serum ACE concentrations has a genetic association in the region, H_2_ = only colorectal cancer risk has a genetic association in the region, H_3_ = both serum ACE concentrations and colorectal cancer risk are associated but have different causal variants, H_4_= both serum ACE concentrations and colorectal cancer risk are associated and share a single causal variant

Supplementary Table 5. Association between genetically-proxied inhibition ACE inhibition and previously reported risk factors for colorectal cancer

| **Previously reported risk factor** | **N** | **Effect estimate**  **(95% CI)** | ***P*-value** |
| --- | --- | --- | --- |
| Body mass index (SD change) | 461,460 | -0.018 (-0.042, 0.007) | 0.11 |
| Low density-lipoprotein cholesterol (mg/dL), SD | 9,961 | 0.050 (-0.024, 0.124) | 0.18 |
| Serum total cholesterol (SD) | 21,491 | -0.040 (-0.109, 0.029) | 0.23 |
| Serum iron (SD, µmol/L) | 23,896 | -0.020 (-0.094, 0.054) | 0.61 |
| Insulin-like growth factor-1 (Nmol/L) | 9,732 | 3.297 (-1.131, 7.726) | 0.15 |

Effect represents the unit change in colorectal cancer risk factor per genetically proxied inhibition of ACE equivalent to a 1 mmHg decrease in systolic blood pressure.

For analyses of genetically-proxied ACE inhibition and low-density lipoprotein cholesterol, two SNPs (rs12452187, rs11655956) were not available and 1 SNP (rs118138685) was removed because of palindromic alleles with ambiguous effect allele frequencies not permitting strands to be matched. in the LDL-C respective outcome dataset and no proxy SNP was available. For analyses of iron, 1 SNP (rs11655956) was not included because of palindromic alleles with ambiguous effect allele frequencies. For analyses of insulin-like growth factor 1, 2 SNPs (rs11655956, rs118138685) were not included because of palindromic alleles with ambiguous effect allele frequencies.

Supplementary Table 6. Association between genetically-proxied ACE inhibition and colorectal cancer risk in iterative leave-one-out analysis

| **SNP removed** | **OR (95% CI)** | ***P*-value** |
| --- | --- | --- |
| rs11650201 | 1.13 (1.05-1.22) | 8.6 x 10^-4^ |
| rs11655956 | 1.14 (1.06-1.22) | 2.7 x 10^-4^ |
| rs118121655 | 1.14 (1.06-1.23) | 5.3 x 10^-4^ |
| rs12452187 | 1.13 (1.05-1.22) | 8.6 x 10^-4^ |
| rs12709437 | 1.13 (1.05-1.22) | 1.1 x 10^-3^ |
| rs141118688 | 1.13 (1.05-1.22) | 8.6 x 10^-4^ |
| rs28656895 | 1.14 (1.07-1.22) | 6.3 x 10^-5^ |
| rs3730025 | 1.14 (1.06-1.22) | 2.7 x 10^-4^ |
| rs4343 | 1.12 (1.02-1.23) | 1.8 x 10^-2^ |
| rs4365 | 1.13 (1.05-1.22) | 1.1 x 10^-3^ |
| rs4968780 | 1.13 (1.05-1.21) | 6.1 x 10^-4^ |
| rs79480822 | 1.14 (1.06-1.22) | 6.7 x 10^-4^ |
| rs80311894 | 1.14 (1.06-1.22) | 6.7 x 10^-4^ |

SNP = Single-Nucleotide Polymorphism. OR represents the exponential change in odds of cancer per genetically proxied inhibition of ACE equivalent to a 1 mmHg decrease in systolic blood pressure.

Supplementary Table 7. Posterior probabilities under differing hypotheses relating the associations between systolic blood pressure (in *ADRB1*) and small cell lung carcinoma risk

| **Configuration** | **H_0_** | **H_1_** | **H_2_** | **H_3_** | **H_4_** |
| --- | --- | --- | --- | --- | --- |
|  | 5.11 x 10^-36^ | 0.73 | 1.76 x 10^-36^ | 0.25 | 1.53 x 10^-2^ |

H_0_ = neither systolic blood pressure (in *ADRB1*) nor small cell lung carcinoma risk has a genetic association in the region, H_1_ = only systolic blood pressure (in *ADRB1*) has a genetic association in the region, H_2_ = only small cell lung carcinoma risk has a genetic association in the region, H_3_ = both systolic blood pressure (in *ADRB1*) and small cell lung carcinoma risk are associated but have different causal variants, H_4_= both systolic blood pressure (in *ADRB1*) and small cell lung carcinoma risk are associated and share a single causal variant

Supplementary Table 8. Association between genetically-proxied ADRB1 inhibition and risk of overall and subtype-specific breast, colorectal, prostate, and lung cancer risk using instrument constructed from a GWAS unadjusted for BMI

| **Outcome** | **N (cases, controls)** | **OR (95% CI)** | ***P*-value** |
| --- | --- | --- | --- |
| Breast cancer | 122,977; 105,974 | 1.04 (1.01-1.07) | 0.02 |
| ER+ Breast cancer | 69,501; 105,974 | 1.04 (1.00-1.08) | 0.06 |
| ER- Breast cancer | 21,468; 105,974 | 1.00 (0.94-1.06) | 0.89 |
| Colorectal cancer | 58,221; 67,694 | 0.99 (0.95-1.04) | 0.71 |
| Colon cancer | 32,002; 64,159 | 1.00 (0.95-1.05) | 0.87 |
| Rectal cancer | 16,212; 64,159 | 1.02 (0.95-1.08) | 0.67 |
| Lung cancer | 29,863; 55,586 | 1.01 (0.94-1.08) | 0.87 |
| Lung adenocarcinoma | 11,245; 54,619 | 0.97 (0.89-1.05) | 0.41 |
| Small cell lung carcinoma | 2,791; 20,580 | 0.88 (0.77-1.02) | 0.08 |
| Squamous cell lung cancer | 7,704; 54,763 | 0.98 (0.89-1.07) | 0.63 |
| Prostate cancer | 79,148; 61,106 | 0.99 (0.95-1.03) | 0.70 |
| Advanced prostate cancer | 15,167; 58,308 | 0.97 (0.91-1.04) | 0.43 |

OR represents the exponential change in odds of cancer per genetically proxied inhibition of ADRB1 equivalent to a 1 mmHg decrease in systolic blood pressure.

Supplementary Table 9. Posterior probabilities under differing hypotheses relating the associations between systolic blood pressure (in *SLC12A3*) and ER- breast cancer risk

| **Configuration** | **H_0_** | **H_1_** | **H_2_** | **H_3_** | **H_4_** |
| --- | --- | --- | --- | --- | --- |
|  | 4.33 x 10^-2^ | 0.82 | 3.95 x 10^-3^ | 7.5 x 10^-2^ | 5.56 x 10^-2^ |

H_0_ = neither systolic blood pressure (in *SLC12A3*) nor ER- breast cancer risk has a genetic association in the region, H_1_ = only systolic blood pressure (in *SLC12A3*) has a genetic association in the region, H_2_ = only ER- breast cancer risk has a genetic association in the region, H_3_ = both systolic blood pressure (in *SLC12A3*) and ER- breast cancer risk are associated but have different causal variants, H_4_= both systolic blood pressure (in *SLC12A3*) and ER- breast cancer risk are associated and share a single causal variant

Supplementary Table 10. Posterior probabilities under differing hypotheses relating the associations between colon ACE gene expression and colorectal cancer risk

| **Configuration** | **H_0_** | **H_1_** | **H_2_** | **H_3_** | **H_4_** |
| --- | --- | --- | --- | --- | --- |
|  | 4.74 x 10^-257^ | 0.68 | 1.69 x 10^-257^ | 0.24 | 7.64 x 10^-2^ |

H_0_ = neither colon ACE concentrations nor colorectal cancer risk has a genetic association in the region, H_1_ = only colon ACE concentrations has a genetic association in the region, H_2_ = only colorectal cancer risk has a genetic association in the region, H_3_ = both colon ACE concentrations and colorectal cancer risk are associated but have different causal variants, H_4_= both colon ACE concentrations and colorectal cancer risk are associated and share a single causal variant

**The PRACTICAL Consortium**

[**http://practical.icr.ac.uk/**](http://practical.icr.ac.uk/)

Rosalind A. Eeles^1,2^, Christopher A. Haiman^3^, Zsofia Kote-Jarai^1^, Fredrick R. Schumacher^4,5^, Sara Benlloch^6,1^, Ali Amin Al Olama^6,7^, Kenneth Muir^8,9^, Sonja I. Berndt^10^, David V. Conti^3^, Fredrik Wiklund^11^, Stephen Chanock^10^, Ying Wang^12^, Victoria L. Stevens^12^, Catherine M. Tangen^13^, Jyotsna Batra^14,15^, Judith A. Clements^14,15^, APCB BioResource (Australian Prostate Cancer BioResource)^14,15^, Henrik Grönberg^11^, Nora Pashayan^16,17^, Johanna Schleutker^18,19^, Demetrius Albanes^10^, Stephanie Weinstein^10^, Alicja Wolk^20^, Catharine M. L. West^21^, Lorelei A. Mucci^22^, Géraldine Cancel-Tassin^23,24^, Stella Koutros^10^, Karina Dalsgaard Sørensen^25,26^, Eli Marie Grindedal^27^, David E. Neal^28,29,30^, Freddie C. Hamdy^31,32^, Jenny L. Donovan^33^, Ruth C. Travis^34^, Robert J. Hamilton^35,36^, Sue Ann Ingles^37^, Barry S. Rosenstein^38,39^, Yong-Jie Lu^40^, Graham G. Giles^41,42,43^, Adam S. Kibel^44^, Ana Vega^45,46,47^, Manolis Kogevinas^48,49,50,51^, Kathryn L. Penney^52^, Jong Y. Park^53^, Janet L. Stanford^54,55^, Cezary Cybulski^56^, Børge G. Nordestgaard^57,58^, Sune F. Nielsen^57,58^, Hermann Brenner^59,60,61^, Christiane Maier^62^, Jeri Kim^63^, Esther M. John^64^, Manuel R. Teixeira^65,66,67^, Susan L. Neuhausen^68^, Kim De Ruyck^69^, Azad Razack^70^, Lisa F. Newcomb^54,71^, Davor Lessel^72^, Radka Kaneva^73^, Nawaid Usmani^74,75^, Frank Claessens^76^, Paul A. Townsend^77,78^, Manuela Gago-Dominguez^79,80^, Monique J. Roobol^81^, Florence Menegaux^82^, Kay-Tee Khaw^83^, Lisa Cannon-Albright^84,85^, Hardev Pandha^78^, Stephen N. Thibodeau^86^, David J. Hunter^87^, Peter Kraft^88^, William J. Blot^89,90^, Elio Riboli^91^

^1^The Institute of Cancer Research, London, SM2 5NG, UK
^2^Royal Marsden NHS Foundation Trust, London, SW3 6JJ, UK
^3^Center for Genetic Epidemiology, Department of Preventive Medicine, Keck School of Medicine, University of Southern California/Norris Comprehensive Cancer Center, Los Angeles, CA 90015, USA
^4^Department of Population and Quantitative Health Sciences, Case Western Reserve University, Cleveland, OH 44106-7219, USA
^5^Seidman Cancer Center, University Hospitals, Cleveland, OH 44106, USA.
^6^Centre for Cancer Genetic Epidemiology, Department of Public Health and Primary Care, University of Cambridge, Strangeways Research Laboratory, Cambridge CB1 8RN, UK
^7^University of Cambridge, Department of Clinical Neurosciences, Stroke Research Group, R3, Box 83, Cambridge Biomedical Campus, Cambridge CB2 0QQ, UK
^8^Division of Population Health, Health Services Research and Primary Care, University of Manchester, Oxford Road, Manchester, M13 9PL, UK
^9^Warwick Medical School, University of Warwick, Coventry, CV4 7AL, UK
^10^Division of Cancer Epidemiology and Genetics, National Cancer Institute, NIH, Bethesda, Maryland, 20892, USA
^11^Department of Medical Epidemiology and Biostatistics, Karolinska Institute, SE-171 77 Stockholm, Sweden
^12^Department of Population Science, American Cancer Society, 250 Williams Street, Atlanta, GA 30303, USA
^13^SWOG Statistical Center, Fred Hutchinson Cancer Research Center, Seattle, WA 98109, USA
^14^Australian Prostate Cancer Research Centre-Qld, Institute of Health and Biomedical Innovation and School of Biomedical Sciences, Queensland University of Technology, Brisbane QLD 4059, Australia
^15^Translational Research Institute, Brisbane, Queensland 4102, Australia
^16^Department of Applied Health Research, University College London, London, WC1E 7HB, UK
^17^Centre for Cancer Genetic Epidemiology, Department of Oncology, University of Cambridge, Strangeways Laboratory, Worts Causeway, Cambridge, CB1 8RN, UK
^18^Institute of Biomedicine, University of Turku, Finland
^19^Department of Medical Genetics, Genomics, Laboratory Division, Turku University Hospital, PO Box 52, 20521 Turku, Finland
^20^Department of Surgical Sciences, Uppsala University, 75185 Uppsala, Sweden
^21^Division of Cancer Sciences, University of Manchester, Manchester Academic Health Science Centre, Radiotherapy Related Research, The Christie Hospital NHS Foundation Trust, Manchester, M13 9PL UK
^22^Department of Epidemiology, Harvard T. H. Chan School of Public Health, Boston, MA 02115, USA
^23^CeRePP, Tenon Hospital, F-75020 Paris, France.
^24^Sorbonne Universite, GRC n°5 , AP-HP, Tenon Hospital, 4 rue de la Chine, F-75020 Paris, France
^25^Department of Molecular Medicine, Aarhus University Hospital, Palle Juul-Jensen Boulevard 99, 8200 Aarhus N, Denmark
^26^Department of Clinical Medicine, Aarhus University, DK-8200 Aarhus N
^27^Department of Medical Genetics, Oslo University Hospital, 0424 Oslo, Norway
^28^Nuffield Department of Surgical Sciences, University of Oxford, Room 6603, Level 6, John Radcliffe Hospital, Headley Way, Headington, Oxford, OX3 9DU, UK
^29^University of Cambridge, Department of Oncology, Box 279, Addenbrooke's Hospital, Hills Road, Cambridge CB2 0QQ, UK
^30^Cancer Research UK, Cambridge Research Institute, Li Ka Shing Centre, Cambridge, CB2 0RE, UK
^31^Nuffield Department of Surgical Sciences, University of Oxford, Oxford, OX1 2JD, UK
^32^Faculty of Medical Science, University of Oxford, John Radcliffe Hospital, Oxford, UK
^33^Population Health Sciences, Bristol Medical School, University of Bristol, BS8 2PS, UK
^34^Cancer Epidemiology Unit, Nuffield Department of Population Health, University of Oxford, Oxford, OX3 7LF, UK
^35^Dept. of Surgical Oncology, Princess Margaret Cancer Centre, Toronto ON M5G 2M9, Canada
^36^Dept. of Surgery (Urology), University of Toronto, Canada
^37^Department of Preventive Medicine, Keck School of Medicine, University of Southern California/Norris Comprehensive Cancer Center, Los Angeles, CA 90015, USA
^38^Department of Radiation Oncology and Department of Genetics and Genomic Sciences, Box 1236, Icahn School of Medicine at Mount Sinai, One Gustave L. Levy Place, New York, NY 10029, USA
^39^Department of Genetics and Genomic Sciences, Icahn School of Medicine at Mount Sinai, New York, NY 10029-5674 , USA.
^40^Centre for Cancer Biomarker and Biotherapeutics, Barts Cancer Institute, Queen Mary University of London, John Vane Science Centre, Charterhouse Square, London, EC1M 6BQ, UK
^41^Cancer Epidemiology Division, Cancer Council Victoria, 615 St Kilda Road, Melbourne, VIC 3004, Australia
^42^Centre for Epidemiology and Biostatistics, Melbourne School of Population and Global Health, The University of Melbourne, Grattan Street, Parkville, VIC 3010, Australia
^43^Precision Medicine, School of Clinical Sciences at Monash Health, Monash University, Clayton, Victoria 3168, Australia
^44^Division of Urologic Surgery, Brigham and Womens Hospital, 75 Francis Street, Boston, MA 02115, USA
^45^Fundación Pública Galega Medicina Xenómica, Santiago de Compostela, 15706, Spain.
^46^Instituto de Investigación Sanitaria de Santiago de Compostela, Santiago De Compostela, 15706, Spain.
^47^Centro de Investigación en Red de Enfermedades Raras (CIBERER), Spain
^48^ISGlobal, Barcelona, Spain
^49^IMIM (Hospital del Mar Medical Research Institute), Barcelona, Spain
^50^CIBER Epidemiología y Salud Pública (CIBERESP), 28029 Madrid, Spain
^51^Universitat Pompeu Fabra (UPF), Barcelona, Spain
^52^Channing Division of Network Medicine, Department of Medicine, Brigham and Women's Hospital/Harvard Medical School, Boston, MA 02115, USA
^53^Department of Cancer Epidemiology, Moffitt Cancer Center, 12902 Magnolia Drive, Tampa, FL 33612, USA
^54^Division of Public Health Sciences, Fred Hutchinson Cancer Research Center, Seattle, Washington, 98109-1024, USA
^55^Department of Epidemiology, School of Public Health, University of Washington, Seattle, Washington 98195, USA
^56^International Hereditary Cancer Center, Department of Genetics and Pathology, Pomeranian Medical University, 70-115 Szczecin, Poland
^57^Faculty of Health and Medical Sciences, University of Copenhagen, 2200 Copenhagen, Denmark
^58^Department of Clinical Biochemistry, Herlev and Gentofte Hospital, Copenhagen University Hospital, Herlev, 2200 Copenhagen, Denmark
^59^Division of Clinical Epidemiology and Aging Research, German Cancer Research Center (DKFZ), D-69120, Heidelberg, Germany
^60^German Cancer Consortium (DKTK), German Cancer Research Center (DKFZ), D-69120 Heidelberg, Germany
^61^Division of Preventive Oncology, German Cancer Research Center (DKFZ) and National Center for Tumor Diseases (NCT), Im Neuenheimer Feld 460, 69120 Heidelberg, Germany
^62^Humangenetik Tuebingen, Paul-Ehrlich-Str 23, D-72076 Tuebingen, Germany
^63^The University of Texas M. D. Anderson Cancer Center, Department of Genitourinary Medical Oncology, 1515 Holcombe Blvd., Houston, TX 77030, USA
^64^Departments of Epidemiology & Population Health and of Medicine, Division of Oncology, Stanford Cancer Institute, Stanford University School of Medicine, Stanford, CA 94304 USA
^65^Department of Genetics, Portuguese Oncology Institute of Porto (IPO-Porto), 4200-072 Porto, Portugal
^66^Biomedical Sciences Institute (ICBAS), University of Porto, 4050-313 Porto, Portugal
^67^Cancer Genetics Group, IPO-Porto Research Center (CI-IPOP), Portuguese Oncology Institute of Porto (IPO-Porto), 4200-072 Porto, Portugal
^68^Department of Population Sciences, Beckman Research Institute of the City of Hope, 1500 East Duarte Road, Duarte, CA 91010, 626-256-HOPE (4673)
^69^Ghent University, Faculty of Medicine and Health Sciences, Basic Medical Sciences, Proeftuinstraat 86, B-9000 Gent
^70^Department of Surgery, Faculty of Medicine, University of Malaya, 50603 Kuala Lumpur, Malaysia
^71^Department of Urology, University of Washington, 1959 NE Pacific Street, Box 356510, Seattle, WA 98195, USA
^72^Institute of Human Genetics, University Medical Center Hamburg-Eppendorf, D-20246 Hamburg, Germany
^73^Molecular Medicine Center, Department of Medical Chemistry and Biochemistry, Medical University of Sofia, Sofia, 2 Zdrave Str., 1431 Sofia, Bulgaria
^74^Department of Oncology, Cross Cancer Institute, University of Alberta, 11560 University Avenue, Edmonton, Alberta, Canada T6G 1Z2
^75^Division of Radiation Oncology, Cross Cancer Institute, 11560 University Avenue, Edmonton, Alberta, Canada T6G 1Z2
^76^Molecular Endocrinology Laboratory, Department of Cellular and Molecular Medicine, KU Leuven, BE-3000, Belgium
^77^Division of Cancer Sciences, Manchester Cancer Research Centre, Faculty of Biology, Medicine and Health, Manchester Academic Health Science Centre, NIHR Manchester Biomedical Research Centre, Health Innovation Manchester, Univeristy of Manchester, M13 9WL
^78^The University of Surrey, Guildford, Surrey, GU2 7XH, UK
^79^Genomic Medicine Group, Galician Foundation of Genomic Medicine, Instituto de Investigacion Sanitaria de Santiago de Compostela (IDIS), Complejo Hospitalario Universitario de Santiago, Servicio Galego de Saúde, SERGAS, 15706, Santiago de Compostela, Spai
^80^University of California San Diego, Moores Cancer Center, Department of Family Medicine and Public Health, University of California San Diego, La Jolla, CA 92093-0012, USA
^81^Department of Urology, Erasmus University Medical Center, 3015 CE Rotterdam, The Netherlands
^82^"Exposome and Heredity", CESP (UMR 1018), Faculté de Médecine, Université Paris-Saclay, Inserm, Gustave Roussy, Villejuif
^83^Clinical Gerontology Unit, University of Cambridge, Cambridge, CB2 2QQ, UK
^84^Division of Epidemiology, Department of Internal Medicine, University of Utah School of Medicine, Salt Lake City, Utah 84132, USA
^85^George E. Wahlen Department of Veterans Affairs Medical Center, Salt Lake City, Utah 84148, USA
^86^Department of Laboratory Medicine and Pathology, Mayo Clinic, Rochester, MN 55905, USA
^87^Nuffield Department of Population Health, University of Oxford, United Kingdom
^88^Program in Genetic Epidemiology and Statistical Genetics, Department of Epidemiology, Harvard School of Public Health, Boston, MA, USA
^89^Division of Epidemiology, Department of Medicine, Vanderbilt University Medical Center, 2525 West End Avenue, Suite 800, Nashville, TN 37232 USA.
^90^International Epidemiology Institute, Rockville, MD 20850, USA
^91^Department of Epidemiology and Biostatistics, School of Public Health, Imperial College London, SW7 2AZ, UK

**MEGASTROKE consortium**

Rainer Malik ^1^, Ganesh Chauhan ^2^, Matthew Traylor ^3^, Muralidharan Sargurupremraj ^4,5^, Yukinori Okada ^6,7,8^, Aniket Mishra ^4,5^, Loes Rutten-Jacobs ^3^, Anne-Katrin Giese ^9^, Sander W van der Laan ^10^, Solveig Gretarsdottir ^11^, Christopher D Anderson ^12,13,14,14^, Michael Chong ^15^, Hieab HH Adams ^16,17^, Tetsuro Ago ^18^, Peter Almgren ^19^, Philippe Amouyel ^20,21^, Hakan Ay ^22,13^, Traci M Bartz ^23^, Oscar R Benavente ^24^, Steve Bevan ^25^, Giorgio B Boncoraglio ^26^, Robert D Brown, Jr.  ^27^, Adam S Butterworth ^28,29^, Caty Carrera ^30,31^, Cara L Carty ^32,33^, Daniel I Chasman ^34,35^, Wei-Min Chen ^36^, John W Cole ^37^, Adolfo Correa ^38^, Ioana Cotlarciuc ^39^, Carlos Cruchaga ^40,41^, John Danesh ^28,42,43,44^, Paul IW de Bakker ^45,46^, Anita L DeStefano ^47,48^, Marcel den Hoed ^49^, Qing Duan ^50^, Stefan T Engelter ^51,52^, Guido J Falcone ^53,54^, Rebecca F Gottesman ^55^, Raji P Grewal ^56^, Vilmundur Gudnason ^57,58^, Stefan Gustafsson ^59^, Jeffrey Haessler ^60^, Tamara B Harris ^61^, Ahamad Hassan ^62^, Aki S Havulinna ^63,64^, Susan R Heckbert ^65^, Elizabeth G Holliday ^66,67^, George Howard ^68^, Fang-Chi Hsu ^69^, Hyacinth I Hyacinth ^70^, M Arfan Ikram ^16^, Erik Ingelsson ^71,72^, Marguerite R Irvin ^73^, Xueqiu Jian ^74^, Jordi Jiménez-Conde ^75^, Julie A Johnson ^76,77^, J Wouter Jukema ^78^, Masahiro Kanai ^6,7,79^, Keith L Keene ^80,81^, Brett M Kissela ^82^, Dawn O Kleindorfer ^82^, Charles Kooperberg ^60^, Michiaki Kubo ^83^, Leslie A Lange ^84^, Carl D Langefeld ^85^, Claudia Langenberg ^86^, Lenore J Launer ^87^, Jin-Moo Lee ^88^, Robin Lemmens ^89,90^, Didier Leys ^91^, Cathryn M Lewis ^92,93^, Wei-Yu Lin ^28,94^, Arne G Lindgren ^95,96^, Erik Lorentzen ^97^, Patrik K Magnusson ^98^, Jane Maguire ^99^, Ani Manichaikul ^36^, Patrick F McArdle ^100^, James F Meschia ^101^, Braxton D Mitchell ^100,102^, Thomas H Mosley ^103,104^, Michael A Nalls ^105,106^, Toshiharu Ninomiya ^107^, Martin J O'Donnell ^15,108^, Bruce M Psaty ^109,110,111,112^, Sara L Pulit ^113,45^, Kristiina Rannikmäe ^114,115^, Alexander P Reiner ^65,116^, Kathryn M Rexrode ^117^, Kenneth Rice ^118^, Stephen S Rich ^36^, Paul M Ridker ^34,35^, Natalia S Rost ^9,13^, Peter M Rothwell ^119^, Jerome I Rotter ^120,121^, Tatjana Rundek ^122^, Ralph L Sacco ^122^, Saori Sakaue ^7,123^, Michele M Sale ^124^, Veikko Salomaa ^63^, Bishwa R Sapkota ^125^, Reinhold Schmidt ^126^, Carsten O Schmidt  ^127^, Ulf Schminke ^128^, Pankaj Sharma ^39^, Agnieszka Slowik ^129^, Cathie LM Sudlow ^114,115^, Christian Tanislav ^130^, Turgut Tatlisumak ^131,132^, Kent D Taylor ^120,121^, Vincent NS Thijs ^133,134^, Gudmar Thorleifsson ^11^, Unnur Thorsteinsdottir ^11^, Steffen Tiedt ^1^, Stella Trompet ^135^, Christophe Tzourio ^5,136,137^, Cornelia M van Duijn ^138,139^, Matthew Walters ^140^, Nicholas J Wareham ^86^, Sylvia Wassertheil-Smoller ^141^, James G Wilson ^142^, Kerri L Wiggins ^109^, Qiong Yang ^47^, Salim Yusuf ^15^, Najaf Amin ^16^, Hugo S Aparicio ^185,48^, Donna K Arnett ^186^, John Attia ^187^, Alexa S Beiser ^47,48^, Claudine Berr ^188^, Julie E Buring ^34,35^, Mariana Bustamante ^189^, Valeria Caso ^190^, Yu-Ching Cheng ^191^, Seung Hoan Choi ^192,48^, Ayesha Chowhan ^185,48^, Natalia Cullell ^31^, Jean-François Dartigues ^193,194^, Hossein Delavaran ^95,96^, Pilar Delgado ^195^, Marcus Dörr ^196,197^, Gunnar Engström ^19^, Ian Ford ^198^, Wander S Gurpreet ^199^, Anders Hamsten ^200,201^, Laura Heitsch ^202^, Atsushi Hozawa ^203^, Laura Ibanez ^204^, Andreea Ilinca ^95,96^, Martin Ingelsson ^205^, Motoki Iwasaki ^206^, Rebecca D Jackson ^207^, Katarina Jood ^208^, Pekka Jousilahti ^63^, Sara Kaffashian ^4,5^, Lalit Kalra ^209^, Masahiro Kamouchi ^210^, Takanari Kitazono ^211^, Olafur Kjartansson ^212^, Manja Kloss ^213^, Peter J Koudstaal ^214^, Jerzy Krupinski ^215^, Daniel L Labovitz ^216^, Cathy C Laurie ^118^, Christopher R Levi ^217^, Linxin Li ^218^, Lars Lind ^219^, Cecilia M Lindgren ^220,221^, Vasileios Lioutas ^222,48^, Yong Mei Liu ^223^, Oscar L Lopez ^224^, Hirata Makoto ^225^, Nicolas Martinez-Majander ^172^, Koichi Matsuda ^225^, Naoko Minegishi ^203^, Joan Montaner  ^226^, Andrew P Morris ^227,228^, Elena Muiño ^31^, Martina Müller-Nurasyid ^229,230,231^, Bo Norrving ^95,96^, Soichi Ogishima ^203^, Eugenio A Parati ^232^, Leema Reddy Peddareddygari ^56^, Nancy L Pedersen ^98,233^, Joanna Pera ^129^, Markus Perola ^63,234^, Alessandro Pezzini ^235^, Silvana Pileggi ^236^, Raquel Rabionet ^237^, Iolanda Riba-Llena ^30^, Marta Ribasés ^238^, Jose R Romero ^185,48^, Jaume Roquer ^239,240^, Anthony G Rudd ^241,242^, Antti-Pekka Sarin ^243,244^, Ralhan Sarju ^199^, Chloe Sarnowski ^47,48^, Makoto Sasaki ^245^, Claudia L Satizabal ^185,48^, Mamoru Satoh ^245^, Naveed Sattar ^246^, Norie Sawada ^206^, Gerli Sibolt ^172^, Ásgeir Sigurdsson ^247^, Albert Smith ^248^, Kenji Sobue ^245^, Carolina Soriano-Tárraga ^240^, Tara Stanne ^249^, O Colin Stine ^250^, David J Stott ^251^, Konstantin Strauch ^229,252^, Takako Takai  ^203^, Hideo Tanaka ^253,254^, Kozo Tanno ^245^, Alexander Teumer ^255^, Liisa Tomppo ^172^, Nuria P Torres-Aguila ^31^, Emmanuel Touze ^256,257^, Shoichiro Tsugane  ^206^, Andre G Uitterlinden ^258^, Einar M Valdimarsson ^259^, Sven J van der Lee ^16^, Henry Völzke ^255^, Kenji Wakai  ^253^, David Weir ^260^, Stephen R Williams ^261^, Charles DA Wolfe ^241,242^, Quenna Wong ^118^, Huichun Xu ^191^, Taiki Yamaji ^206^, Dharambir K Sanghera ^125,169,170^, Olle Melander ^19^, Christina Jern ^171^, Daniel Strbian ^172,173^, Israel Fernandez-Cadenas ^31,30^, W T Longstreth, Jr ^174,65^, Arndt Rolfs ^175^, Jun Hata ^107^, Daniel Woo ^82^, Jonathan Rosand ^12,13,14^, Guillaume Pare ^15^, Jemma C Hopewell ^176^, Danish Saleheen ^177^, Kari Stefansson ^11,178^, Bradford B Worrall ^179^, Steven J Kittner ^37^, Sudha Seshadri ^180,48^, Myriam Fornage ^74,181^, Hugh S Markus ^3^, Joanna MM Howson ^28^, Yoichiro Kamatani ^6,182^, Stephanie Debette ^4,5^, Martin Dichgans ^1,183,184^

1 Institute for Stroke and Dementia Research (ISD), University Hospital, LMU Munich, Munich, Germany

2 Centre for Brain Research, Indian Institute of Science, Bangalore, India

3 Stroke Research Group, Division of Clinical Neurosciences, University of Cambridge, UK

4 INSERM U1219 Bordeaux Population Health Research Center, Bordeaux, France

5 University of Bordeaux, Bordeaux, France

6 Laboratory for Statistical Analysis, RIKEN Center for Integrative Medical Sciences, Yokohama, Japan

7 Department of Statistical Genetics, Osaka University Graduate School of Medicine, Osaka, Japan

8 Laboratory of Statistical Immunology, Immunology Frontier Research Center (WPI-IFReC), Osaka University, Suita, Japan.

9 Department of Neurology, Massachusetts General Hospital, Harvard Medical School, Boston, MA, USA

10 Laboratory of Experimental Cardiology, Division of Heart and Lungs, University Medical Center Utrecht, University of Utrecht, Utrecht,Netherlands

11 deCODE genetics/AMGEN inc, Reykjavik, Iceland

12 Center for Genomic Medicine, Massachusetts General Hospital (MGH), Boston, MA, USA

13 J. Philip Kistler Stroke Research Center, Department of Neurology, MGH, Boston, MA, USA

14 Program in Medical and Population Genetics, Broad Institute, Cambridge, MA, USA

15 Population Health Research Institute, McMaster University, Hamilton, Canada

16 Department of Epidemiology, Erasmus University Medical Center, Rotterdam, Netherlands

17 Department of Radiology and Nuclear Medicine, Erasmus University Medical Center, Rotterdam, Netherlands

18 Department of Medicine and Clinical Science, Graduate School of Medical Sciences, Kyushu University, Fukuoka, Japan

19 Department of Clinical Sciences, Lund University, Malmö, Sweden

20 Univ. Lille, Inserm, Institut Pasteur de Lille, LabEx DISTALZ-UMR1167, Risk factors and molecular determinants of aging-related diseases, F-59000 Lille, France

21 Centre Hosp. Univ Lille, Epidemiology and Public Health Department, F-59000 Lille, France

22 AA Martinos Center for Biomedical Imaging, Department of Radiology, Massachusetts General Hospital, Harvard Medical School, Boston, MA, USA

23 Cardiovascular Health Research Unit, Departments of Biostatistics and Medicine, University of Washington, Seattle, WA, USA

24 Division of Neurology, Faculty of Medicine, Brain Research Center, University of British Columbia, Vancouver, Canada

25 School of Life Science, University of Lincoln, Lincoln, UK

26 Department of Cerebrovascular Diseases, Fondazione IRCCS Istituto Neurologico "Carlo Besta", Milano, Italy

27 Department of Neurology, Mayo Clinic Rochester, Rochester, MN, USA

28 MRC/BHF Cardiovascular Epidemiology Unit, Department of Public Health and Primary Care, University of Cambridge, Cambridge, UK

29 The National Institute for Health Research Blood and Transplant Research Unit in Donor Health and Genomics, University of Cambridge, UK

30 Neurovascular Research Laboratory, Vall d'Hebron Institut of Research, Neurology and Medicine Departments-Universitat Autònoma de Barcelona, Vall d’Hebrón Hospital, Barcelona, Spain

31 Stroke Pharmacogenomics and Genetics, Fundacio Docència i Recerca MutuaTerrassa, Terrassa, Spain

32 Children's Research Institute, Children's National Medical Center, Washington, DC, USA

33 Center for Translational Science, George Washington University, Washington, DC, USA

34 Division of Preventive Medicine, Brigham and Women's Hospital, Boston, MA, USA

35 Harvard Medical School, Boston, MA, USA

36 Center for Public Health Genomics, Department of Public Health Sciences, University of Virginia, Charlottesville, VA, USA

37 Department of Neurology, University of Maryland School of Medicine and Baltimore VAMC, Baltimore, MD, USA

38 Departments of Medicine, Pediatrics and Population Health Science, University of Mississippi Medical Center, Jackson, MS, USA

39 Institute of Cardiovascular Research, Royal Holloway University of London, UK  &  Ashford and St Peters Hospital, Surrey UK

40 Department of Psychiatry,The Hope Center Program on Protein Aggregation and Neurodegeneration (HPAN),Washington University, School of Medicine, St. Louis, MO, USA

41 Department of Developmental Biology, Washington University School of Medicine, St. Louis, MO, USA

42 NIHR Blood and Transplant Research Unit in Donor Health and Genomics, Department of Public Health and Primary Care, University of Cambridge, Cambridge, UK

43 Wellcome Trust Sanger Institute, Wellcome Trust Genome Campus, Hinxton,  Cambridge, UK

44 British Heart Foundation, Cambridge Centre of Excellence, Department of Medicine, University of Cambridge, Cambridge, UK

45 Department of Medical Genetics, University Medical Center Utrecht, Utrecht, Netherlands

46 Department of Epidemiology, Julius Center for Health Sciences and Primary Care, University Medical Center Utrecht, Utrecht, Netherlands

47 Boston University School of Public Health, Boston, MA, USA

48 Framingham Heart Study, Framingham, MA, USA

49 Department of Immunology, Genetics and Pathology and Science for Life Laboratory, Uppsala University, Uppsala, Sweden

50 Department of Genetics, University of North Carolina, Chapel Hill, NC, USA

51 Department of Neurology and Stroke Center, Basel University Hospital, Switzerland

52 Neurorehabilitation Unit, University and University Center for Medicine of Aging and Rehabilitation Basel, Felix Platter Hospital, Basel, Switzerland

53 Department of Neurology, Yale University School of Medicine, New Haven, CT, USA

54 Program in Medical and Population Genetics, The Broad Institute of Harvard and MIT, Cambridge, MA, USA

55 Department of Neurology, Johns Hopkins University School of Medicine, Baltimore, MD, USA

56 Neuroscience Institute, SF Medical Center, Trenton, NJ, USA

57 Icelandic Heart Association Research Institute, Kopavogur, Iceland

58 University of Iceland, Faculty of Medicine, Reykjavik, Iceland

59 Department of Medical Sciences, Molecular Epidemiology and Science for Life Laboratory, Uppsala University, Uppsala, Sweden

60 Division of Public Health Sciences, Fred Hutchinson Cancer Research Center, Seattle, WA, USA

61 Laboratory of Epidemiology and Population Science, National Institute on Aging, National Institutes of Health, Bethesda, MD, USA

62 Department of Neurology, Leeds General Infirmary, Leeds Teaching Hospitals NHS Trust, Leeds, UK

63 National Institute for Health and Welfare, Helsinki, Finland

64 FIMM - Institute for Molecular Medicine Finland, Helsinki, Finland

65 Department of Epidemiology, University of Washington, Seattle, WA, USA

66 Public Health Stream, Hunter Medical Research Institute, New Lambton, Australia

67 Faculty of Health and Medicine, University of Newcastle, Newcastle, Australia

68 School of Public Health, University of Alabama at Birmingham, Birmingham, AL, USA

69 Department of Biostatistical Sciences, Wake Forest School of Medicine, Winston-Salem, NC, USA

70 Aflac Cancer and Blood Disorder Center, Department of Pediatrics, Emory University School of Medicine, Atlanta, GA, USA

71 Department of Medicine, Division of Cardiovascular Medicine, Stanford University School of Medicine, CA, USA

72 Department of Medical Sciences, Molecular Epidemiology and Science for Life Laboratory, Uppsala University, Uppsala, Sweden

73 Epidemiology, School of Public Health, University of Alabama at Birmingham, USA

74 Brown Foundation Institute of Molecular Medicine, University of Texas Health Science Center at Houston, Houston, TX, USA

75 Neurovascular Research Group (NEUVAS), Neurology Department, Institut Hospital del Mar d'Investigació Mèdica, Universitat Autònoma de Barcelona, Barcelona, Spain

76 Department of Pharmacotherapy and Translational Research and Center for Pharmacogenomics, University of Florida, College of Pharmacy, Gainesville, FL, USA

77 Division of Cardiovascular Medicine, College of Medicine, University of Florida, Gainesville, FL, USA

78 Department of Cardiology, Leiden University Medical Center, Leiden, the Netherlands

79 Program in Bioinformatics and Integrative Genomics, Harvard Medical School, Boston, MA, USA

80 Department of Biology, East Carolina University, Greenville, NC, USA

81 Center for Health Disparities, East Carolina University, Greenville, NC, USA

82 University of Cincinnati College of Medicine, Cincinnati, OH, USA

83 RIKEN Center for Integrative Medical Sciences, Yokohama, Japan

84 Department of Medicine, University of Colorado Denver, Anschutz Medical Campus, Aurora, CO, USA

85 Center for Public Health Genomics and Department of Biostatistical Sciences, Wake Forest School of Medicine, Winston-Salem, NC, USA

86 MRC Epidemiology Unit, University of Cambridge School of Clinical Medicine, Institute of Metabolic Science, Cambridge Biomedical Campus, Cambridge, UK

87 Intramural Research Program, National Institute on Aging, National Institutes of Health, Bethesda, MD, USA

88 Department of Neurology, Radiology, and Biomedical Engineering, Washington University School of Medicine, St. Louis, MO, USA

89 KU Leuven – University of Leuven, Department of Neurosciences,  Experimental Neurology, Leuven, Belgium

90 VIB Center for Brain & Disease Research, University Hospitals Leuven, Department of Neurology, Leuven, Belgium

91 Univ.-Lille, INSERM U 1171. CHU Lille. Lille, France

92 Department of Medical and Molecular Genetics, King's College London, London, UK

93 SGDP Centre, Institute of Psychiatry, Psychology & Neuroscience, King's College London, London, UK

94 Northern Institute for Cancer Research, Paul O'Gorman Building, Newcastle University, Newcastle, UK

95 Department of Clinical Sciences Lund, Neurology, Lund University, Lund, Sweden

96 Department of Neurology and Rehabilitation Medicine, Skåne University Hospital, Lund, Sweden

97 Bioinformatics Core Facility, University of Gothenburg, Gothenburg, Sweden

98 Department of Medical Epidemiology and Biostatistics, Karolinska Institutet, Stockholm, Sweden

99 University of Technology Sydney, Faculty of Health, Ultimo, Australia

100 Department of Medicine, University of Maryland School of Medicine, MD, USA

101 Department of Neurology, Mayo Clinic, Jacksonville, FL, USA

102 Geriatrics Research and Education Clinical Center, Baltimore Veterans Administration Medical Center, Baltimore, MD, USA

103 Division of Geriatrics, School of Medicine, University of Mississippi Medical Center, Jackson, MS, USA

104 Memory Impairment and Neurodegenerative Dementia Center, University of Mississippi Medical Center, Jackson, MS, USA

105 Laboratory of Neurogenetics, National Institute on Aging, National institutes of Health, Bethesda, MD, USA

106 Data Tecnica International, Glen Echo MD, USA

107 Department of Epidemiology and Public Health, Graduate School of Medical Sciences, Kyushu University, Fukuoka, Japan

108 Clinical Research Facility, Department of Medicine, NUI Galway, Galway, Ireland

109 Cardiovascular Health Research Unit, Department of Medicine, University of Washington, Seattle, WA, USA

110 Department of Epidemiology, University of Washington, Seattle, WA

111 Department of Health Services, University of Washington, Seattle, WA, USA

112 Kaiser Permanente Washington Health Research Institute, Seattle, WA, USA

113 Brain Center Rudolf Magnus, Department of Neurology, University Medical Center Utrecht, Utrecht, The Netherlands

114 Usher Institute of Population Health Sciences and Informatics, University of Edinburgh, Edinburgh, UK

115 Centre for Clinical Brain Sciences, University of Edinburgh, Edinburgh, UK

116 Fred Hutchinson Cancer Research Center, University of Washington, Seattle, WA, USA

117 Department of Medicine, Brigham and Women's Hospital, Boston, MA, USA

118 Department of Biostatistics, University of Washington, Seattle, WA, USA

119 Nuffield Department of Clinical Neurosciences, University of Oxford, UK

120 Institute for Translational Genomics and Population Sciences, Los Angeles Biomedical Research Institute at  Harbor-UCLA Medical Center, Torrance, CA, USA

121 Division of Genomic Outcomes, Department of Pediatrics, Harbor-UCLA Medical Center, Torrance, CA, USA

122 Department of Neurology, Miller School of Medicine, University of Miami, Miami, FL, USA

123 Department of Allergy and Rheumatology, Graduate School of Medicine, the University of Tokyo, Tokyo, Japan

124 Center for Public Health Genomics, University of Virginia, Charlottesville, VA, USA

125 Department of Pediatrics, College of Medicine, University of Oklahoma Health Sciences Center, Oklahoma City, OK, USA

126 Department of Neurology, Medical University of Graz, Graz, Austria

127 University Medicine  Greifswald, Institute for Community Medicine, SHIP-KEF, Greifswald, Germany

128 University Medicine  Greifswald,  Department of Neurology, Greifswald, Germany

129 Department of Neurology, Jagiellonian University, Krakow, Poland

130 Department of Neurology, Justus Liebig University, Giessen, Germany

131 Department of Clinical Neurosciences/Neurology, Institute of Neuroscience and Physiology, Sahlgrenska Academy at University of Gothenburg, Gothenburg, Sweden

132 Sahlgrenska University Hospital, Gothenburg, Sweden

133 Stroke Division, Florey Institute of Neuroscience and Mental Health, University of Melbourne, Heidelberg, Australia

134 Austin Health, Department of Neurology, Heidelberg, Australia

135 Department of Internal Medicine, Section Gerontology and Geriatrics, Leiden University Medical Center, Leiden, the Netherlands

136 INSERM U1219, Bordeaux, France

137 Department of Public Health, Bordeaux University Hospital, Bordeaux, France

138 Genetic Epidemiology Unit, Department of Epidemiology, Erasmus University Medical Center Rotterdam, Netherlands

139 Center for Medical Systems Biology, Leiden, Netherlands

140 School of Medicine, Dentistry and Nursing at the University of Glasgow, Glasgow, UK

141 Department of Epidemiology and Population Health, Albert Einstein College of Medicine, NY, USA

142 Department of Physiology and Biophysics, University of Mississippi Medical Center, Jackson, MS, USA

143 A full list of members and affiliations appears in the Supplementary Note

144 Department of Human Genetics, McGill University, Montreal, Canada

145 Department of Pathophysiology, Institute of Biomedicine and Translation Medicine, University of Tartu, Tartu, Estonia

146 Department of Cardiac Surgery, Tartu University Hospital, Tartu, Estonia

147 Clinical Gene Networks AB,Stockholm, Sweden

148 Department of Genetics and Genomic Sciences, The Icahn Institute for Genomics and Multiscale Biology Icahn School of Medicine at Mount Sinai, New York, NY , USA

149 Department of Pathophysiology, Institute of Biomedicine and Translation Medicine, University of Tartu, Biomeedikum, Tartu, Estonia

150 Integrated Cardio Metabolic Centre, Department of Medicine, Karolinska Institutet, Karolinska Universitetssjukhuset, Huddinge, Sweden.

151 Clinical Gene Networks AB, Stockholm, Sweden

152 Sorbonne Universités, UPMC Univ. Paris 06, INSERM, UMR_S 1166, Team Genomics & Pathophysiology of Cardiovascular Diseases, Paris, France

153 ICAN Institute for Cardiometabolism and Nutrition, Paris, France

154 Department of Biomedical Engineering, University of Virginia, Charlottesville, VA, USA

155 Group Health Research Institute, Group Health Cooperative, Seattle, WA, USA

156 Seattle Epidemiologic Research and Information Center, VA Office of Research and Development, Seattle, WA, USA

157 Cardiovascular Research Center, Massachusetts General Hospital, Boston, MA, USA

158 Department of Medical Research, Bærum Hospital, Vestre Viken Hospital Trust, Gjettum, Norway

159 Saw Swee Hock School of Public Health, National University of Singapore and National University Health System, Singapore

160 National Heart and Lung Institute, Imperial College London, London, UK

161 Department of Gene Diagnostics and Therapeutics, Research Institute, National Center for Global Health and Medicine, Tokyo, Japan

162 Department of Epidemiology, Tulane University School of Public Health and Tropical Medicine, New Orleans, LA, USA

163 Department of Cardiology,University Medical Center Groningen, University of Groningen, Netherlands

164 MRC-PHE Centre for Environment and Health, School of Public Health, Department of Epidemiology and Biostatistics, Imperial College London, London, UK

165 Department of Epidemiology and Biostatistics, Imperial College London, London, UK

166 Department of Cardiology, Ealing Hospital NHS Trust, Southall, UK

167 National Heart, Lung and Blood Research Institute, Division of Intramural Research, Population Sciences Branch, Framingham, MA, USA

168 A full list of members and affiliations appears at the end of the manuscript

169 Department of Phamaceutical Sciences, Collge of Pharmacy, University of Oklahoma Health Sciences Center, Oklahoma City, OK, USA

170 Oklahoma Center for Neuroscience, Oklahoma City, OK, USA

171 Department of Pathology and Genetics, Institute of Biomedicine, The Sahlgrenska Academy at University of Gothenburg, Gothenburg, Sweden

172 Department of Neurology, Helsinki University Hospital, Helsinki, Finland

173 Clinical Neurosciences, Neurology, University of Helsinki, Helsinki, Finland

174 Department of Neurology, University of Washington, Seattle, WA, USA

175 Albrecht Kossel Institute, University Clinic of Rostock, Rostock, Germany

176 Clinical Trial Service Unit and Epidemiological Studies Unit, Nuffield Department of Population Health, University of Oxford, Oxford, UK

177 Department of Genetics, Perelman School of Medicine, University of Pennsylvania, PA, USA

178 Faculty of Medicine, University of Iceland, Reykjavik, Iceland

179 Departments of Neurology and Public Health Sciences, University of Virginia School of Medicine, Charlottesville, VA, USA

180 Department of Neurology, Boston University School of Medicine, Boston, MA, USA

181 Human Genetics Center, University of Texas Health Science Center at Houston, Houston, TX, USA

182 Center for Genomic Medicine, Kyoto University Graduate School of Medicine, Kyoto, Japan

183 Munich Cluster for Systems Neurology (SyNergy), Munich, Germany

184 German Center for Neurodegenerative Diseases (DZNE), Munich, Germany

185 Boston University School of Medicine, Boston, MA, USA

186 University of Kentucky College of Public Health, Lexington, KY, USA

187 University of Newcastle and Hunter Medical Research Institute, New Lambton, Australia

188 Univ. Montpellier, Inserm, U1061, Montpellier, France

189 Centre for Research in Environmental Epidemiology, Barcelona, Spain

190 Department of Neurology, Università degli Studi di Perugia, Umbria, Italy

191 Department of Medicine, University of Maryland School of Medicine, Baltimore, MD, USA

192 Broad Institute, Cambridge, MA, USA

193 Univ. Bordeaux, Inserm, Bordeaux Population Health Research Center, UMR 1219, Bordeaux, France

194 Bordeaux University Hospital, Department of Neurology, Memory Clinic, Bordeaux, France

195 Neurovascular Research Laboratory. Vall d'Hebron Institut of Research, Neurology and Medicine Departments-Universitat Autònoma de Barcelona. Vall d’Hebrón Hospital, Barcelona, Spain

196 University Medicine Greifswald, Department of Internal Medicine B, Greifswald, Germany

197 DZHK, Greifswald, Germany

198 Robertson Center for Biostatistics, University of Glasgow, Glasgow, UK

199 Hero DMC Heart Institute, Dayanand Medical College & Hospital, Ludhiana, India

200 Atherosclerosis Research Unit, Department of Medicine Solna, Karolinska Institutet, Stockholm, Sweden

201 Karolinska Institutet, Stockholm, Sweden

202 Division of Emergency Medicine, and Department of Neurology, Washington University School of Medicine, St. Louis, MO, USA

203 Tohoku Medical Megabank Organization, Sendai, Japan

204 Department of Psychiatry, Washington University School of Medicine, St. Louis, MO, USA

205 Department of Public Health and Caring Sciences / Geriatrics, Uppsala University, Uppsala, Sweden

206 Epidemiology and Prevention Group, Center for Public Health Sciences, National Cancer Center, Tokyo, Japan

207 Department of Internal Medicine and the Center for Clinical and Translational Science, The Ohio State University, Columbus, OH, USA

208 Institute of Neuroscience and Physiology, the Sahlgrenska Academy at University of Gothenburg, Goteborg, Sweden

209 Department of Basic and Clinical Neurosciences, King's College London, London, UK

210 Department of Health Care Administration and Management, Graduate School of Medical Sciences, Kyushu University, Japan

211 Department of Medicine and Clinical Science, Graduate School of Medical Sciences, Kyushu University, Japan

212 Landspitali National University Hospital, Departments of Neurology & Radiology, Reykjavik, Iceland

213 Department of Neurology, Heidelberg University Hospital, Germany

214 Department of Neurology, Erasmus University Medical Center

215 Hospital Universitari Mutua Terrassa, Terrassa (Barcelona), Spain

216 Albert Einstein College of Medicine, Montefiore Medical Center, New York, NY, USA

217 John Hunter Hospital, Hunter Medical Research Institute and University of Newcastle, Newcastle, NSW, Australia

218 Centre for Prevention of Stroke and Dementia, Nuffield Department of Clinical Neurosciences, University of Oxford, UK

219 Department of Medical Sciences, Uppsala University, Uppsala, Sweden

220 Genetic and Genomic Epidemiology Unit, Wellcome Trust Centre for Human Genetics, University of Oxford, Oxford, UK

221 The Wellcome Trust Centre for Human Genetics, Oxford, UK

222 Beth Israel Deaconess Medical Center, Boston, MA, USA

223 Wake Forest School of Medicine, Wake Forest, NC, USA

224 Department of Neurology, University of Pittsburgh, Pittsburgh, PA, USA

225 BioBank Japan, Laboratory of Clinical Sequencing, Department of Computational biology and medical Sciences, Graduate school of Frontier Sciences, The University of Tokyo, Tokyo, Japan

226 Neurovascular Research Laboratory, Vall d'Hebron Institut of Research, Neurology and Medicine Departments-Universitat Autònoma de Barcelona. Vall d’Hebrón Hospital, Barcelona, Spain

227 Department of Biostatistics, University of Liverpool, Liverpool, UK

228 Wellcome Trust Centre for Human Genetics, University of Oxford, Oxford, UK

229 Institute of Genetic Epidemiology, Helmholtz Zentrum München - German Research Center for Environmental Health, Neuherberg, Germany

230 Department of Medicine I, Ludwig-Maximilians-Universität, Munich, Germany

231 DZHK (German Centre for Cardiovascular Research), partner site Munich Heart Alliance, Munich, Germany

232 Department of Cerebrovascular Diseases, Fondazione IRCCS Istituto Neurologico “Carlo Besta”, Milano, Italy

233 Karolinska Institutet, MEB, Stockholm, Sweden

234 University of Tartu, Estonian Genome Center, Tartu, Estonia, Tartu, Estonia

235 Department of Clinical and Experimental Sciences, Neurology Clinic, University of Brescia, Italy

236 Translational Genomics Unit, Department of Oncology, IRCCS Istituto di Ricerche Farmacologiche Mario Negri, Milano, Italy

237 Department of Genetics, Microbiology and Statistics, University of Barcelona, Barcelona, Spain

238 Psychiatric Genetics Unit, Group of Psychiatry, Mental Health and Addictions, Vall d’Hebron Research Institute (VHIR), Universitat Autònoma de Barcelona, Biomedical Network Research Centre on Mental Health (CIBERSAM), Barcelona, Spain

239 Department of Neurology, IMIM-Hospital del Mar, and Universitat Autònoma de Barcelona, Spain

240 IMIM (Hospital del Mar Medical Research Institute), Barcelona, Spain

241 National Institute for Health Research Comprehensive Biomedical Research Centre, Guy's & St. Thomas' NHS Foundation Trust and King's College London, London, UK

242 Division of Health and Social Care Research, King's College London, London, UK

243 FIMM-Institute for Molecular Medicine Finland, Helsinki, Finland

244 THL-National Institute for Health and Welfare, Helsinki, Finland

245 Iwate Tohoku Medical Megabank Organization, Iwate Medical University, Iwate, Japan

246 BHF Glasgow Cardiovascular Research Centre, Faculty of Medicine, Glasgow, UK

247 deCODE Genetics/Amgen, Inc., Reykjavik, Iceland

248 Icelandic Heart Association, Reykjavik, Iceland

249 Institute of Biomedicine, the Sahlgrenska Academy at University of Gothenburg, Goteborg, Sweden

250 Department of Epidemiology, University of Maryland School of Medicine, Baltimore, MD, USA

251 Institute of Cardiovascular and Medical Sciences, Faculty of Medicine, University of Glasgow, Glasgow, UK

252 Chair of Genetic Epidemiology, IBE, Faculty of Medicine, LMU Munich, Germany

253 Division of Epidemiology and Prevention, Aichi Cancer Center Research Institute, Nagoya, Japan

254 Department of Epidemiology, Nagoya University Graduate School of Medicine, Nagoya, Japan

255 University Medicine Greifswald, Institute for Community Medicine, SHIP-KEF, Greifswald, Germany

256 Department of Neurology, Caen University Hospital, Caen, France

257 University of Caen Normandy, Caen, France

258 Department of Internal Medicine, Erasmus University Medical Center, Rotterdam, Netherlands

259 Landspitali University Hospital, Reykjavik, Iceland

260 Survey Research Center, University of Michigan, Ann Arbor, MI, USA

261 University of Virginia Department of Neurology, Charlottesville, VA, USA

**Supplementary Material citations**

7.	Díez-Obrero, V.e.a., *Genetic effects on transcriptome profiles in colon epithelium provide functional insights for genetic risk loci.* Cell Mol Gastroenterol Hepatol, 2021. **In Press**.

8.	Langfelder, P. and S. Horvath, *WGCNA: an R package for weighted correlation network analysis.* BMC Bioinformatics, 2008. **9**: p. 559.

9.	Watanabe, K., et al., *Functional mapping and annotation of genetic associations with FUMA.* Nat Commun, 2017. **8**(1): p. 1826.
